# Paradoxical Sex-Specific Patterns of Autoantibodies Response to SARS-CoV-2 Infection

**DOI:** 10.1101/2021.07.15.21260603

**Authors:** Yunxian Liu, Joseph E. Ebinger, Rowann Mostafa, Petra Budde, Jana Gajewski, Brian Walker, Sandy Joung, Min Wu, Manuel Bräutigam, Franziska Hesping, Elena Schäfer, Ann-Sophie Schubert, Hans-Dieter Zucht, Jonathan Braun, Gil Y. Melmed, Kimia Sobhani, Moshe Arditi, Jennifer E. Van Eyk, Susan Cheng, Justyna Fert-Bober

**Affiliations:** Department of Cardiology, Smidt Heart Institute, Cedars-Sinai Medical Center, Los Angeles, California, USA; Advanced Clinical Biosystems Institute, Department of Biomedical Sciences, Cedars-Sinai Medical Center, Los Angeles, California, USA; Division of Gastroenterology, Department of Medicine, Cedars-Sinai Medical Center, Los Angeles, California, USA; Department of Pathology and Laboratory Medicine, Cedars-Sinai Medical Center, Los Angeles, CA, USA; Department of Medical Research, Oncimmune Germany GmbH, Dortmund, Germany; Ruhr University Bochum, TU Dortmund University, Germany, Department of Pediatrics, Division of Infectious Diseases and Immunology, Infectious and Immunologic Diseases Research Center (IIDRC), and Department of Biomedical Sciences, Cedars-Sinai Medical Center, Los Angeles, California, USA

**Author notes:** Correspondence. Justyna Fert-Bober, PhD, Department of Cardiology, Smidt Heart Institute, Cedars Sinai Medical Center, Los Angeles, CA; Susan Cheng, MD, MPH, Department of Cardiology, Smidt Heart Institute, Cedars Sinai Medical Center, Los Angeles, CA; Jennifer Van Eyk, PhD, Department of Cardiology, Smidt Heart Institute, Cedars-Sinai Medical Center, Los Angeles, California, USA; phone: (310). Equal contribution. **Conflict of Interest:** PB, JG, MB, FH, ES, ASS, and HDZ work for Oncimmune, a company that performed the serological assays on the biospecimens that were collected for this study. The remaining authors have no competing interests.

**Keywords:** SARS-CoV-2, COVID-19, AABs, Autoantigen selectivity, Sex differences.

## Abstract

**Background:** Amidst the millions of individuals affected directly by the pandemic, pronounced sex differences in the susceptibility and response to SARS-CoV-2 infection remain poorly understood. Emerging evidence has highlighted the potential importance of autoimmune activation in modulating not only the acute response but also recovery trajectories following SARS-CoV-2 exposure. Given that immune-inflammatory activity can be sex-biased in the setting of severe COVID-19 illness, we deliberately examined sex-specific autoimmune reactivity to SARS-CoV-2 in the absence of extreme clinical disease.

**Methods:** We used a bead-based array containing over 90 autoantigens previously linked to a range of classic autoimmune diseases to assess autoantibody (AAB) titers in 177 participants. All participants had confirmed evidence of prior SARS-CoV-2 infection based on presence of positive anti-nucleocapsid IgG serology results (Abbott Diagnostics, Abbott Park, Illinois). We used multivariate analysis to determine whether sex-bias was associated with increased rates of AABs reactivity and symptom burden after SARS-CoV2 infection.

**Results:** 82.4% of AABs reactivity was associated with being male compared to 17.6% with female. We found a diversity of AABs responses that exhibited sex-specific patterns of frequency distribution as well as associations with symptomatology and symptom burden.

**Conclusion:** Our results reveal a remarkable sex-specific prevalence and selectivity of AAB responses to SARS-CoV-2. Further understanding of the nature of triggered and persistent AAB activation among men and women exposed to SARS-CoV-2 will be essential for developing effective interventions against immune-mediated sequelae of COVID-19.

## INTRODUCTION

Mechanisms underlying sex differences in both susceptibility and response to SARS-CoV-2 infection remain poorly understood. Biological sex differences have become manifest with respect to vulnerability to infection, adaptive immune responses, and the equilibrium of inflammation and tissue repair in the resolution of infection (1). Recent evidence points to the possible contributions of triggering and persistence of autoimmune activation in SARS-CoV-2-infected COVID-19 patients (2, 3). Intriguingly, despite classic autoimmune diseases being more prevalent in females, emerging studies have revealed a paradoxical male predominance of autoimmune activation in the setting of severe COVID-19 illness (4). The extent to which such paradoxical sex differences in triggered autoimmunity may exist and persist across the broader clinical spectrum of SARS-CoV-2 infection is unclear. Recognizing that sex bias is potentially introduced when assessing autoimmune activation in the setting of more severe forms of COVID-19 illness, we deliberately aimed to interrogate sex-specific autoimmune activation after SARS-CoV-2 exposure in the absence of any extreme manifestations of clinical disease. Therefore, using an array to detect autoantibodies (AABs) to over 90 antigens previously linked to a range of classic autoimmune diseases, we sought to comprehensively examine the diversity of AAB responses in male and female health care workers (HCWs) who were exposed to SARS-CoV-2 and experienced even minimal or no symptoms.

## RESULTS

Use of the multiplex assay resulted in complete AABs array profiling for all 177 plasma samples collected from our primary HCW participants, in addition to samples from the 53 healthy control and the 6 systemic lupus erythematosus (SLE) patient comparators. Demographic and clinical characteristics of the primary study sample are shown in **Table S2**. The primary cohort (N=177) had a mean age of 35 [IQR: 30-44] years including 65.0% women, 68.4% of non-white race, 28.2% of Hispanic/Latinx ethnicity, and 87% having reported at least mild symptoms associated with COVID-19.

### Sex-Specific Frequency of Symptoms

As shown in **Figure 1**, the vast majority of symptoms related to prior COVID-19 infection were experienced similarly by men and women. Although specific symptoms appeared to be reported more frequently by men (e.g., chills, fever, shortness of breath, diarrhea, conjunctivitis) and other symptoms were reported more frequently by women (e.g., loss of appetite, nausea, and productive cough), these differences were not statistically significant (**Table S2**). Similar to the specific types of symptoms assessed, we observed that varying degreess of total symptom burden were also distributed relatively equally between the sexes (**Figure 1**).

**Figure 1.**
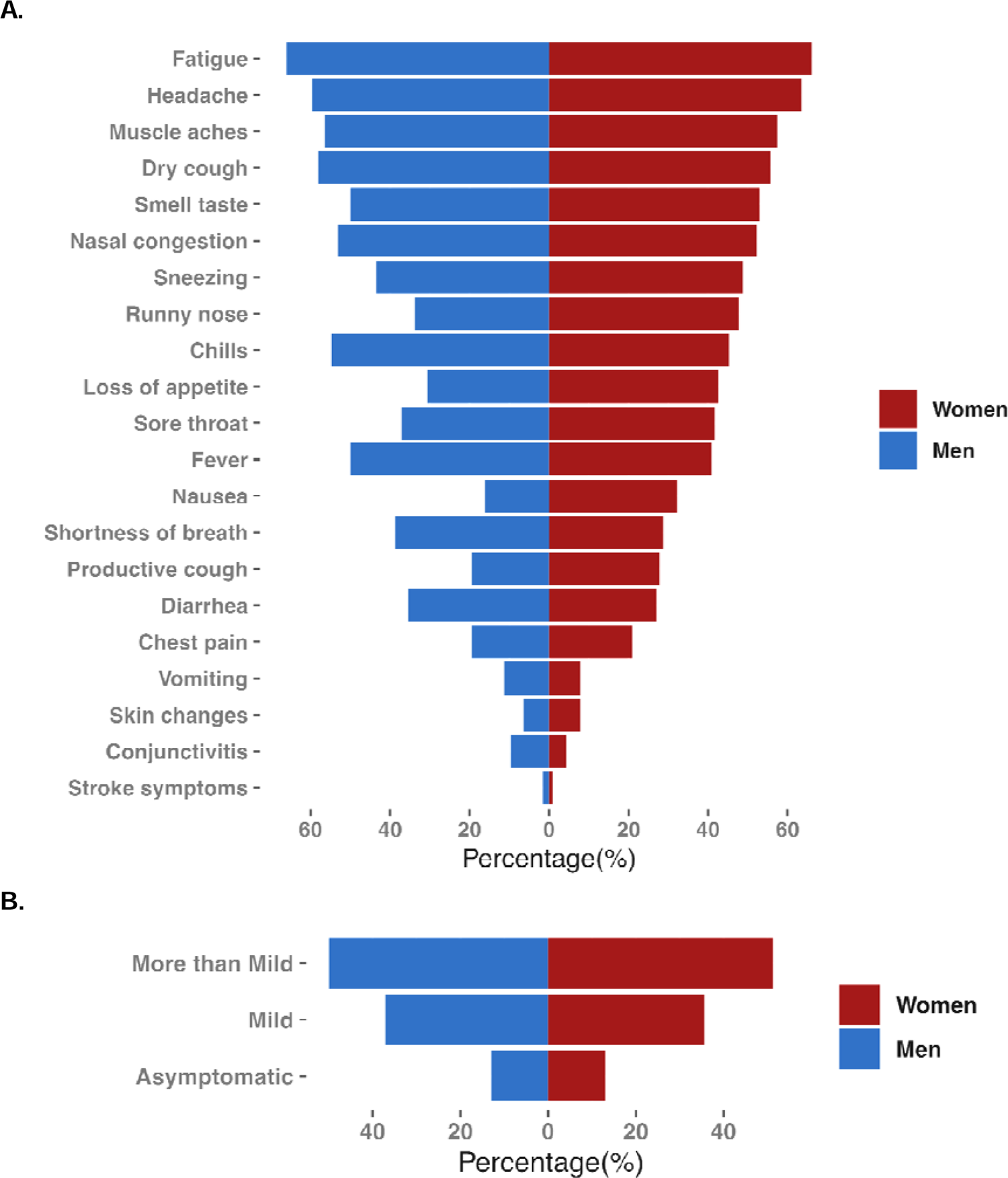
Sex-specific frequency of symptoms type and overall symptoms burden, in men and women previously infected by SARS-CoV-2. In our primary study cohort, the distribution of experienced symptoms was generally similar between men and women (**Panel A**) with some exceptions including certain distinct symptoms being more frequent in men (e.g., chills, fever, conjunctivitis) and other distinct symptoms being more frequent in women (e.g., loss of appetite, nausea). In analyses of overall symptom burden, frequencies of asymptomatic, mildly symptomatic, and more than mildly symptomatic persons were relatively equally distributed between the sexes (**Panel B**).

### Sex-Specific Frequency of AABs Response

In age-adjusted regression analyses, we examined the association of sex (female versus male) with measured plasma levels for each of the 91 autoantibodies assayed. Across the entire cohort, the majority of assayed AABs were associated with male sex and the minority with female sex but the sex-specific frequency and magnitudes of association varied by symptom burden (**Figure 2**). By comparison, a majority of the AABs assayed were significantly associated with SLE compared to healthy controls (HCs), with these associations predominantly seen in females compared to males (**Figure S1**). AABs were also detected in HCs, with trend of AABs reactivity much more pronounce in male however, the AABs that showed reactivity in the HCs group were different compare to AABs reactive in SLE and HCWs individuals (**Figure 2**). In male among HCs AAB reactivity to colony stimulating factor 2 (CSF2), glycoprotein secreted by macrophages, Endothelin converting enzyme 1 (ECE1) and Dihydrolipoamide Branched Chain Transacylase E2 (DBT) involved in protein break down were the most dominated AABs. In female, among HCs reactivity to Transforming growth factor beta (TGFb) and Thyroid Peroxidase (TPO, marker for Hashimoto’s disease or Graves’ disease) showed the highest titer.

**Figure 2.**
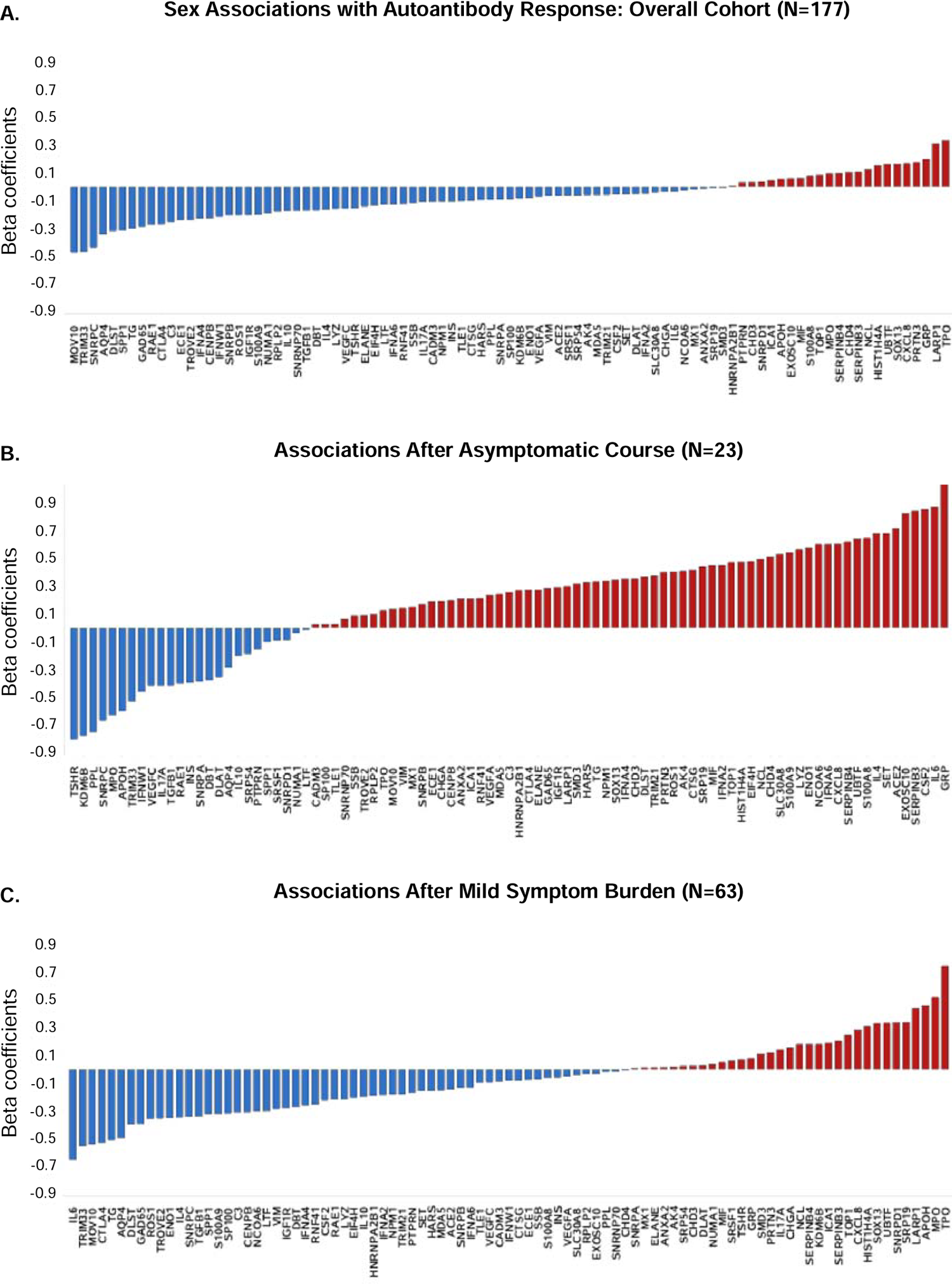
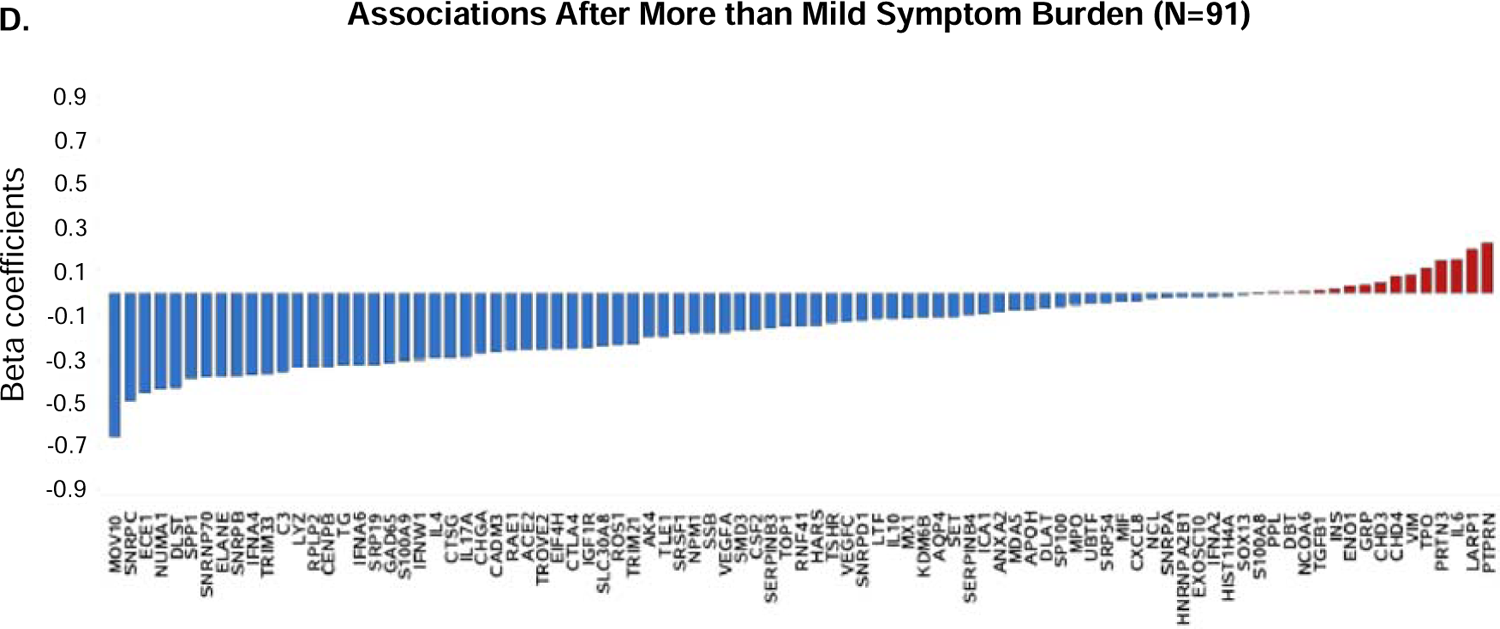
Sex associations with AABs activation by symptoms burden, in men and women previously infected by SARS-CoV-2. The age-adjusted associations of sex (female versus male) with AABs activation across the panel of 91 antigens are shown for the primary cohort overall (**Panel A**) and within persons with varying loads of symptom burden: asymptomatic (**Panel B**), mild symptom burden (**Panel C**), more than mild symptom burden (**Panel D**).

By contrast, among asymptomatic individuals, the breadth and magnitude of AAB reactivity was much more pronounced in females compared to males (**Figure 2B**). Notably, AABs to cytokine and chemokine antigens (IL6 and CSF2) involved in immune defense, together with lung specific proteins (gastrin release peptide (GRP) and serpin family B member 3 (SERPINB3), were predominantly elevated in asymptomatic females. By contrast, thyroid stimulating hormone receptor (TSHR) and lysine demethylase 6B (KDMA6B), which are known primary antigens in autoimmune diseases, were highly expressed in asymptomatic males.

Among all participants who had at least mild symptoms, the range and degrees of AAB reactivity was more pronounced in males compared to females (**Figure 2C-D**). In participants with more than mild symptoms, 82.4% of AABs reactivity was associated with being male compared to 17.6% with female. The most abundant AABs in males including classical nuclear AABs such as small nuclear ribonucleoprotein polypeptide C (SNRPC) (SLE), nuclear mitotic apparatus protein 1 (NUMA1) (systemic sclerosis) and dihydrolipoamide S-succinyltransferase (DLST) (primary biliary cirrhosis). In this setting, the highest expressed antibody was Mov10 RISC complex RNA helicase (MOV10), which has been identified as an AAB that interacts with SARS-CoV-2 proteins (5).

### Sex-Specific Associations of AABs Reactivity with Symptoms

In age-adjusted regression analyses we examined the sex-specific associations of distinct AABs levels with symptomatology (one symptom and marker per model), as shown in **Figures 3-4** and **Tables S3-S6**). In unadjusted regression analyses, 42 of 91 autoantibodies showed statistically significant reactivity that correspond to 18 distinct COVID-19 related symptoms for the whole cohort. In male, 63 out of 91 autoantibodies showed statistically significant reactivity corresponding to 18 distinct COVID-19 related symptoms. In unadjusted analysis in female, 41 out of 91 autoantibodies showed statistically significant reactivity corresponding to 14 COVID-19 related symptoms. After age-adjusted regression analyses, among males, 59 out of 63 AABs were associated with 18 out of all 18 symptoms that demonstrated significant associations. Among females, 38 out of 41 AABs demonstrated significant beta coefficients in relation to 13 out of 14 symptoms (**Figure 3**). Notably, in males, a large number of AABs had increased levels in relation to at least a mild overall symptoms burden while a substantial proportion of these same AABs exhibited lower levels in relation to asymptomatic status. By contrast, a smaller number of AABs exhibited generally consistently increased levels in relation to any level of overall symptom burden in females, including asymptomatic status.

**Figure 3.**
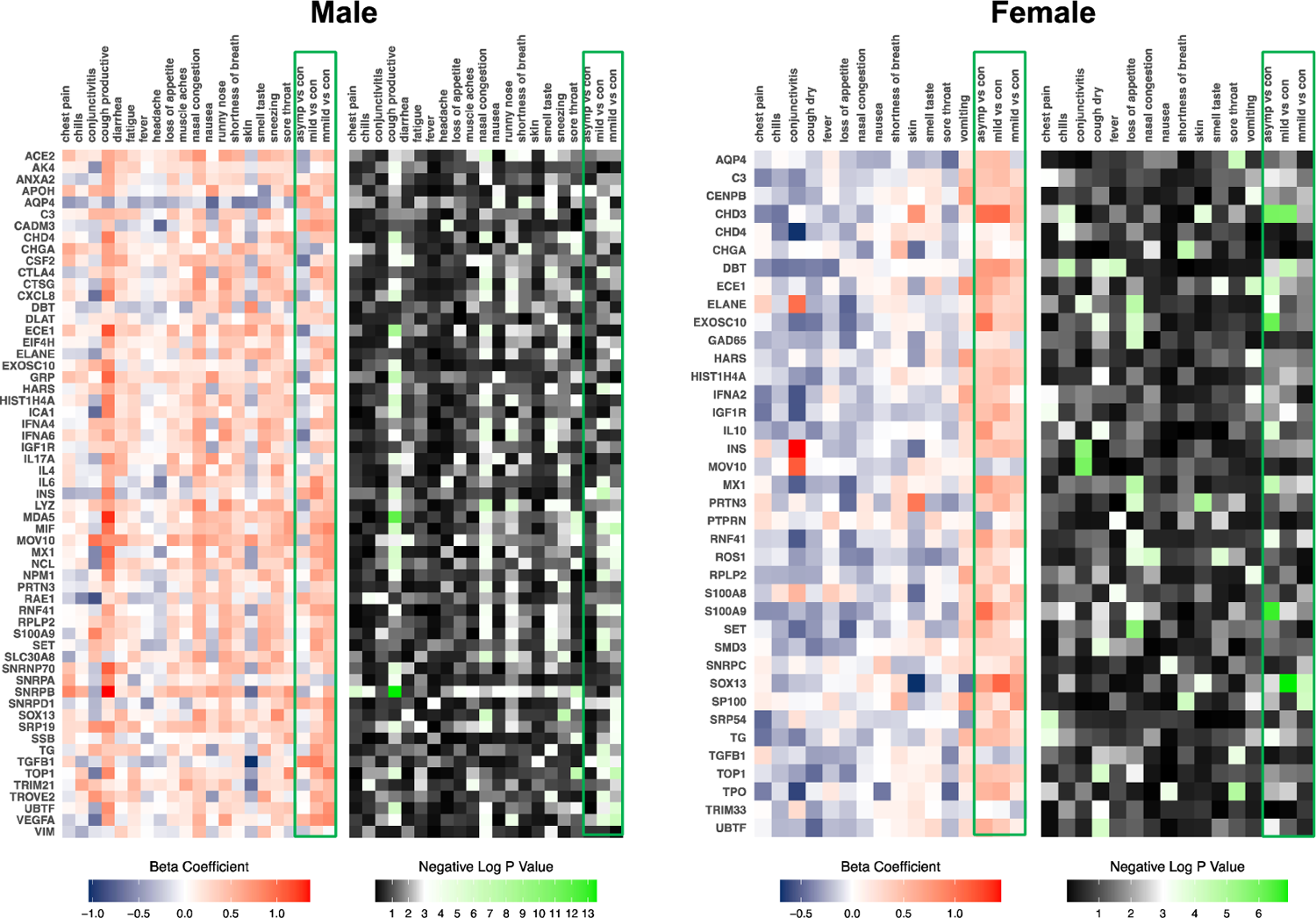
Sex-specific associations of AABs reactivity with symptoms, in persons previously infected by SARS-CoV-2. From age-adjusted regression analyses, beta coefficients and negative log p values were obtained from examining the associations of symptoms with distinct autoantibodies. Associations for men are shown in **Panel A**. Associations for women are shown in **Panel B**.

**Figure 4.**
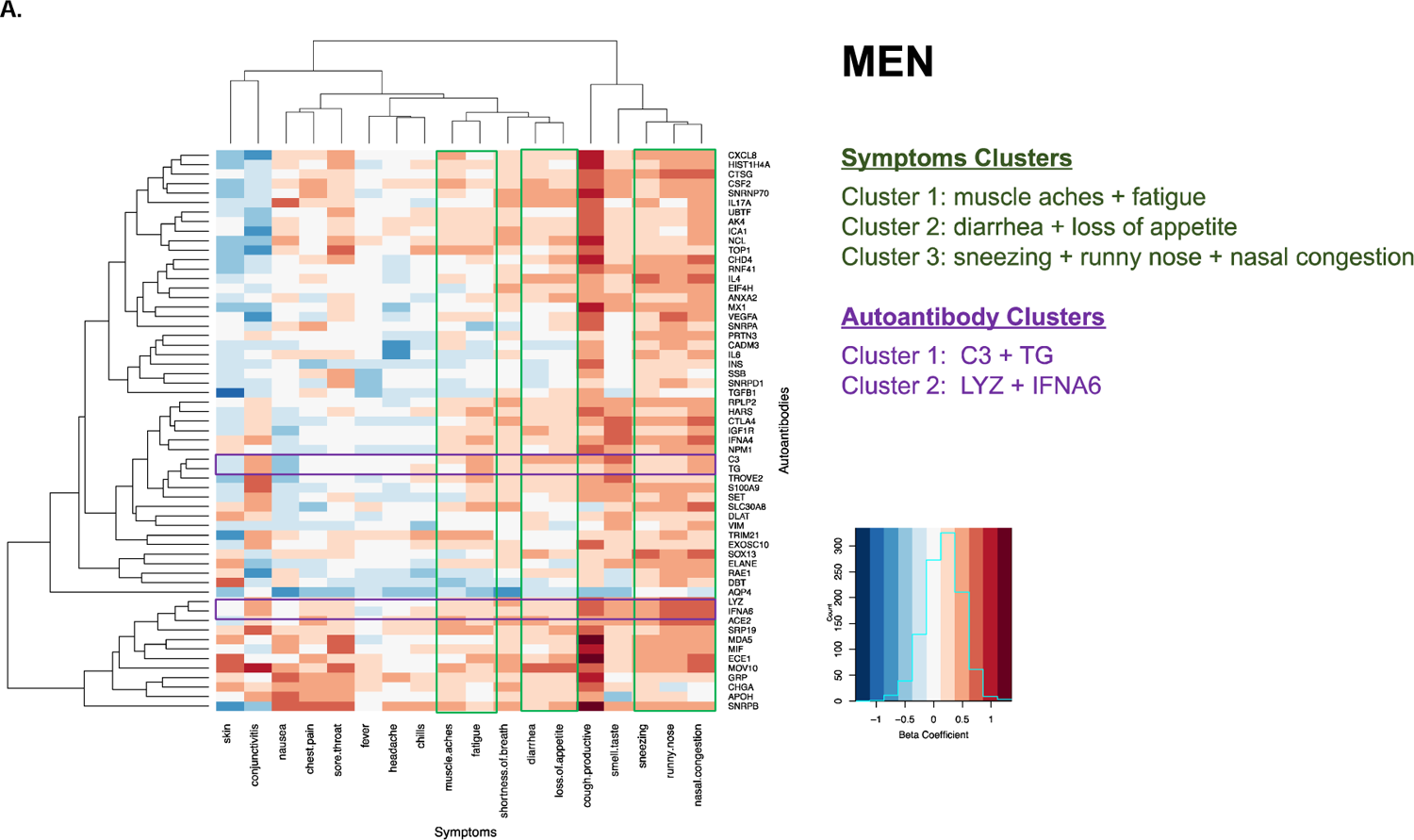
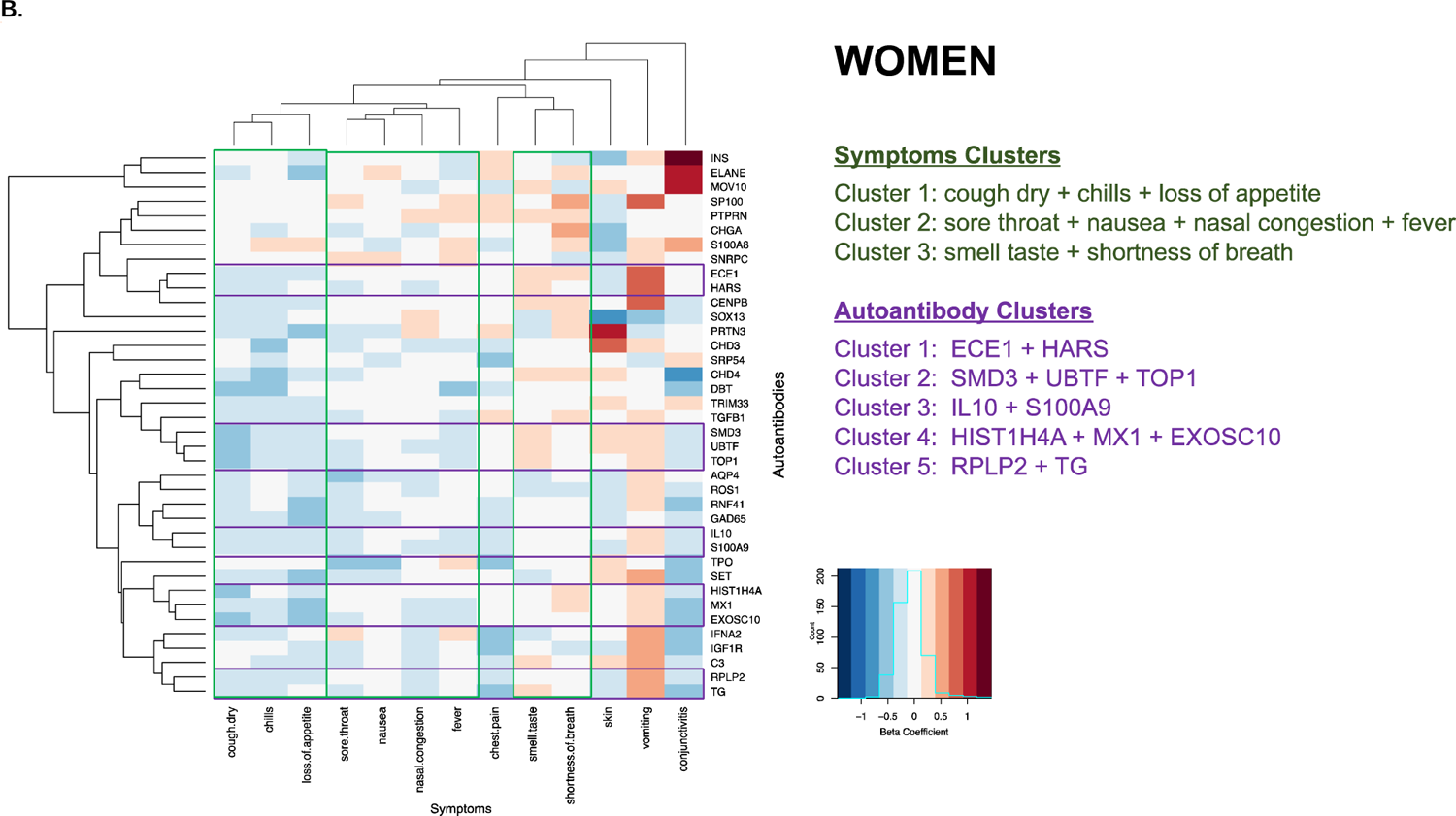
Sex-specific clustering of symptoms and autoantibodies in men and women previously infected with SARS-CoV-2. Symptoms and autoantibodies were grouped based on similar directions and magnitudes of the beta coefficients from age-adjusted regression association analyses, with clusters selected based on a threshold of h=0.5 for autoantibodies and h=1.5 for symptoms from Ward hierarchical clustering. Results are shown for men in **Panel A** and women in **Panel B**.

In males, among the significantly associated AABs, small nuclear ribonucleoprotein polypeptides B (SNRPB), chromodomain helicase DNA binding protein 4 (CHD4) and chromogranin A (CHGA) were the most frequently associated with the distinct symptoms of productive cough and nasal congestion. In females, the AABs to dihydrolipoamide branched chain transacylase E2 (DBT) and ROS proto-oncogene 1, receptor tyrosine kinase (ROS1), were the most frequently elevated in relation to dry cough and loss of appetite (**Figure 3**). Overall profiles of AAB reactivity in relation to symptoms indicated the most frequent and significantly associated AABs in males appeared to follow an SLE-related pattern, whereas AAB profiles in females were more suggestive of poly-autoimmunity (6).

To further investigate the AAB sub-groups and select potential discriminatory symptoms, we applied hierarchical clustering analyses to identify similar magnitudes and directions of associations across AABs. The results of sex-specific two-dimensional clustering of symptoms variables in relation to AABs are shown in **Figure 4**. In males, the initially identified cluster included the symptoms of muscle aches and fatigue (cluster 1), with diarrhea and loss of appetite clustered next (cluster 2), and sneezing, runny nose, and nasal congestion also clustered together (cluster 3). In females, there were also three major clusters identified: dry cough, chilis, and loss of appetite (cluster 1); sore throat, nausea, nasal congestion, and fever (cluster 2); and smell/taste change and shortness of breath (cluster 3). In males, clustering of AABs including C3 with TG antigens (cluster 1) and AABs to antigens representing LYZ and IFNA6 protein (cluster 2). In females, we found 5 clusters in total: ECE1 and HARS (cluster 1); SMD3, UBTF, and TOP1 (cluster 2); IL10 and S100A9 (cluster 3); HIST1H4A, MX1, and EXOSC10 (cluster 4); and, RPLP2 and TG (cluster 5).

## DISCUSSION

In this study, comprehensive profiling of AAB activation in over 170 HCs with prior SARS-CoV-2 infection revealed several important sex-specific findings of interest. First, a surprisingly large number of the diverse autoantibodies assayed were differentially activated in males compared to females. Among previously infected individuals who were asymptomatic, the breadth of AABs response was more prominent in women than in men; by contrast, among previously infected individuals who experienced at least a mild burden of symptoms, the extent of antibody response was far more pronounced in men. Second, we found that the AABs response to symptom clusters were also sex-specific, with certain AABs-symptom associations seen more prominently in men compared to women, across the range of symptom burden. Finally, we observed these sex-specific AABs associations up to 6 months following symptomatology, indicating that SARS-CoV-2 triggers a complement of AABs responses that persists over time – in a sex-specific manner and irrespective of illness severity.

The current study expands from prior work in several ways. Extending from previously studies reporting on presence of post-COVID-19 autoimmunity,(3, 7) we employed a broad array of antibodies to over 90 distinct antigens previously linked to classic autoimmune conditions. Our results reveal a remarkable sex-specific prevalence and selectivity of the AABs response to SARS-CoV-2. Confirming and extending from the findings from prior reports, we found that a majority of our previously infected study participants had detectable circulating AABs against antigens such as ACE2, AQP4, C3, CHD4, CHGA, CXCL8, DBT, ECE1, ELANE, EXOSC10, HARS, HIST1H4A, IGF1R, INS, MOV10, MX1, PRTN3, RNF41, RPLP2, S100A9, SET, SNRPD1, SOX13, TG, TGFB1, TOP1, UBTF. Intriguingly, a distinct set of AABs to 59 antigens were highly correlated with reported symptoms in the male population, while another set of AABs to only 38 antigens were associated with symptoms in females. Notably, in males, we observed AABs associated with symptoms at a high frequency (≥6 symptoms) as well as at a moderate frequency (≥4 symptoms). The high frequency associated AABs included SNRPB, a ribonucleoprotein that is widely prevalent in human SLE (8). The moderate frequency associated AABs included MOV10, CHGA, CHD4, HIST1H4A, ACE2, IFNA6, LYZ, RNF41. Importantly, both MOV10 and IFNA6 have been reported in patients infected with COVID-19 (5, 9). In females, we observed an overall lower frequency of significant symptoms associated AABs when compared to males. The 3 most prominent symptoms in females were associated with AABs to DBT and ROS1. Interestingly, AABs to DBT have been associated with lung cancer (10). Importantly, a number of AABs can be classified as implicated more frequently with systemic disease traits (i.e., multi-organ or multi-system) which may be particularly relevant to the more non-specific symptoms such as fatigue, fever, rashes, cold or allergy-type symptoms, weight loss, and muscular weakness. The sex specificity of triggered AABs reactivity in association with either distinct symptoms, or symptom clusters, may be related not only to sex differences in acute illness but also in post-acute and chronic clinical syndromes experienced by a substantial number of individuals recovering from COVID-19 (11).

While apparently paradoxical at the outset, our sex-specific findings are congruent with ongoing emerging data regarding potential mechanisms underlying sex differences in the susceptibility and response to SARS-CoV-2. Early studies reported that while men and women have similar prevalence, men with COVID-19 are at greater risk for worse outcomes and death independent of age (12, 13). Consistent with these findings, conventional inflammatory markers are founded to be more substantially elevated in men compared to women who are hospitalized for COVID-19 (14). Accordingly, males in our study had a greater than 1.5-fold odds of AABs reactivity after adjusting for age. For classic autoimmune disease, clinical prevalence and incidence of autoimmune diseases tends to follow a male versus female pattern based on pathology. Male-predominant autoimmune diseases usually manifest clinically (i.e., show signs and symptoms of clinical disease) prior to age 50 and are characterized by acute inflammation and a Th1-type response, whereas autoimmune diseases with a greater incidence in females that occur early in life have a clearer antibody-mediated pathology. Autoimmune diseases that have a greater incidence in females and also appear clinically later in life tend to present with evidence of chronic pathology, fibrosis, and increased numbers of autoantibodies are present (15).

The conventional sex bias seen for classic autoimmune diseases has been attributed in part to women who have a generally stronger cellular and humoral immunity, higher levels of circulating antibodies, more numerous circulating CD4+ T cells, and more robust cytokine production in response to immune stressors such as infection (16, 17). By contrast, males are now recognized as more vulnerable to the immune-modulated effects of active SARS-CoV-2 infection likely due to multiple mechanisms (e.g. lower immune cell expression of TLR7, lower observed antibody response, and lower interleukin mediated tissue resilience and tissue repair activity)(17) – and our results demonstrate the persistence of detectable downstream autoimmune sequelae. Importantly, our study also demonstrates for the first time a persistence of autoimmune activation in females compared to males following asymptomatic infection. As context, the stability of AABs in classic autoimmune diseases is known to vary substantially, with some autoantibodies fluctuating with flares of disease, while others remain stable.

We can speculate that the preponderance of AABs positivity in females – in the absence of symptomatic or recognized infection – represents initiation or proliferation and then persistence of self-reactive immunity with implications for post-acute chronic immune-driven disease states. These findings may be particularly relevant to rapidly accumulating evidence of the post-acute SARS-CoV-2 syndromes (e.g. “long-haul COVID”) that can emerge even weeks to months following resolution of mild or asymptomatic infection and with clinical manifestations that appear to differ in women compared to men (11).

The existence of autoantibodies within normal healthy individuals has been already shown by other investigators (18). The fact that across the breadth of AABs assayed in our healthy control sample, titers were also male predominant suggesting that larger population-based screening studies are needed to clarify our understanding of sex differences in basal AAB variation in the absence of clinical disease. Importantly, variations in the AAB titers found in the HCWs were different than those seen in healthy control subjects. In the latter group, the most dominant AAB was for granulocyte-macrophage colony-stimulating factor, also known as colony-stimulating factor 2 (CSF2), well-known to be a regulator of monocyte/macrophage differentiation. By contrast, AAB against CSF2 in the HCWs was barely reactive in the male population and were seen to be upregulated in female in asymptomatic group.

Several limitations of our study merit consideration. Our cohort includes HCWs from a single center who volunteered and responded to surveys, potentially limiting generalizability. We have a relatively small number of male subjects (n=63) that may have limited the ability to detect potential additional predicators of post-COVID autoimmunity; thus, further investigations of larger sized samples are needed. Although this was a prospective study, the survey method involved requesting participants to self-report symptoms occurring up to 6 months prior to the blood draw, contributing to potential recall bias. Whether examined subjectively or objectively, symptomatology can vary not only between but also within individuals over time. Similarly, the status of AAB reactivity may change over time and in relation to the timing of initial or repeated exposures. Thus, future longitudinal studies are warranted to understand temporal trends in similarly measured exposure and outcomes.

In summary, this comprehensive study of AABs to a wide array of antigens found that male sex carries the risk of diverse autoimmune activation following symptomatic COVID-19 illness, whereas female sex carries risk for a distinct profile of autoimmune activation following asymptomatic SARS-CoV-2 exposure. Importantly, both sets of sex-specific AAB reactivity patterns were found to persist up to 6 months following associated symptomatology. Further understanding the nature of triggered and persistent AAB activation among individuals who are exposed to SARS-CoV-2 – and vulnerable to its potentially morbid clinical sequelae – will be essential for developing effective interventions and therapeutics.

## METHODS

### Study Sample

Our primary study cohort was derived from a diverse and unselected population of adults employed in a multisite healthcare delivery system located in Los Angeles County, including individuals with direct patient contact and others with non-patient-oriented work functions. All HCW participants provided detailed survey data on medical history, including prior COVID-19 exposures and infection in addition to COVID-19-related symptoms and exposures (19). For the current study, we included all participants (n=177) who had confirmed evidence of prior SARS-CoV-2 infection based on presence of positive anti-nucleocapsid IgG serology results (Abbott Diagnostics, Abbott Park, Illinois) (19). For all participants, EDTA plasma specimens were transported for assays within 1 hour of phlebotomy to the Cedars-Sinai Department of Pathology and Laboratory Medicine. Our secondary cohorts included healthy controls (n=53) who provided pre-pandemic serum samples obtained from the Bavarian Red Cross (Wiesentheid, Germany) with ethical approval from the Bayerische Landesaerztekammer (Study No. 01/09). We also studied a classic autoimmune disease comparator cohort (n=6) of SLE patients who fulfilled the American College of Rheumatology (ACR) classification criteria for SLE (20) and had pre-pandemic serum samples collected by BioIVT (West Sussex, United Kingdom).

### Autoantibody Assays

We utilized a panel of 91 protein antigens and cytokines in a multiplexed bead-based assay using Luminex FlexMAP 3D technology (21) The selected analytical targets can be grouped into different functional protein families or belong to immune-relevant biological pathways (22). Each antigen was covalently coupled to a distinct magnetic bead region (Luminex Corp, Austin, Texas) by carbodiimide. For each coupling reaction, up to 97 µg antigen and 4 × 105 MagPlex™ beads per color were used (Table S1). Coupling efficiency was confirmed by incubation of 625 beads from each coupled region with a phycoerythrin-conjugated anti-6× HisTag antibody (Abcam, Cambridge, UK) at a concentration of 10 μg/mL for 45 min shaking at 900 rpm and room temperature.

Coupled beads were mixed to a final concentration of 62.5 beads/μL and stored in PBS supplemented with 1% bovine serum albumin, 0.1 % Tween 20 and 0.05% ProClin™ 300 (Merck KGaA, Darmstadt, Germany), at 4 °C until use (8). For analysis, serum or plasma samples were diluted 1:100 in assay buffer (50 % PBS with 1 % BSA, 50% LowCross-Buffer® (Candor Biosciences, Wangen, Germany), 1.3 µg/µl E. coli lysate) and incubated for 20 min at room temperature. Next, the bead’s mix (50 μL) was added to each well of a 96-well plate and incubated with 50 μL of diluted sample (1:100) in assay buffer for 22 h at 4– 8°C in a plate shaker (900 rpm). Subsequently, after washing with PBS/0.1% Tween 20 the beads were incubated with R-phycoerythrin-labelled goat anti-human IgG detection antibody (Ab) (5 µg/ml, Dianova, Hamburg, Germany) for 1 h at RT and washed again. The beads were analyzed in a FlexMap3D instrument (Luminex Corporation, Austin, Texas). The IgG reactivity values were given as median fluorescence intensity (MFI) and antigens fulfilling the minimum bead count criterion (>10 beads measured per bead ID) were exported for data analysis. In addition, triplicates of a COVID-19 positive reference sample (comprised of three HCWs) were run on each plate to calculate median intra-plate coefficient of variation (CV) and median inter-plate CV. The dynamic range was determined using blank samples and a control bead coupled to BSA (control_BSA) for the lower and huIgG (control_huIgGhi) for the upper MFI range. Log MFI of each AAB across all of our study samples (including 177 HCWs, 6 SLEs and 53 HCs) were standardized, by first subtracting the mean, and then dividing by the standard deviation.

### Statistics

Parametric tests and non-parametric tests were used to compare normally distributed continuous variables and non-normally distributed or categorical variables, respectively. Histograms were used to display distribution of symptomatology as well as AABs reactivity against each antigen for the cohort in sex-pooled and sex-specific analyses. Ordinal logistic regression was used to examine the associations between AABs reactivity of each antigen and self-reported symptoms burden, defined as a symptom’s severity score. The symptoms burden score was constructed based on the total number of reported symptoms experienced within 6 months prior to the blood draw wherein a greater number of symptoms corresponded to a higher score (i.e., one point per symptom): we defined asymptomatic as represented by a score of 0, mild symptom burden as a score of 1 to 7, and more than mild symptom burden as a score of >7. All statistical analyses were conducted using R (v3.5.1) and statistical significance was defined as a two-tailed P value <0.05.

### Study approval

All participants provided written informed consent and all protocols were approved by the Cedars-Sinai institutional review board.

## AUTHOR CONTRIBUTION

JFB, JEVE, JB, KS and SC conceived and designed the overall CORALE study. JFB, JB, KS, JEVE, and SC acquired the CORALE data. PB, JG AND H-D Z conceived the AABS analysis of the study, and YL, MW, NS, PB, JG, JFB, KS, JEVE, SC, and MA analyzed the data. JFB, SC, and YL drafted the manuscript, and all authors edited the manuscript.

## Data Availability

Requests for de-identified data may be directed to the corresponding authors (Jennifer E. Van Eyk, Susan Cheng, Justyna Fert-Bober) and will be reviewed by the Office of Research Administration at Cedars-Sinai Medical Center prior to issuance of data sharing agreements. Data limitations are designed to ensure patient and participant confidentiality.

## ACKNOWLEDGEMENTS

We are grateful to all the front-line HCWsin our healthcare system who continue to be dedicated to delivering the highest quality care for all patients. We would like to thank the following people for their collective effort: Koen Raedschelders, PhD, Danica-Mae Manalo, MS, James Stewart, PhD, John Prostko, MS, Edwin Frias, BS from Abbott.

## DATA AVAILABILITY

Requests for de-identified data may be directed to the corresponding authors (JEVE, SC, JFB) and will be reviewed by the Office of Research Administration at Cedars-Sinai Medical Center prior to issuance of data sharing agreements. Data limitations are designed to ensure patient and participant confidentiality.

## SUPPLEMENTAL MATERIAL

**Figure S1.**
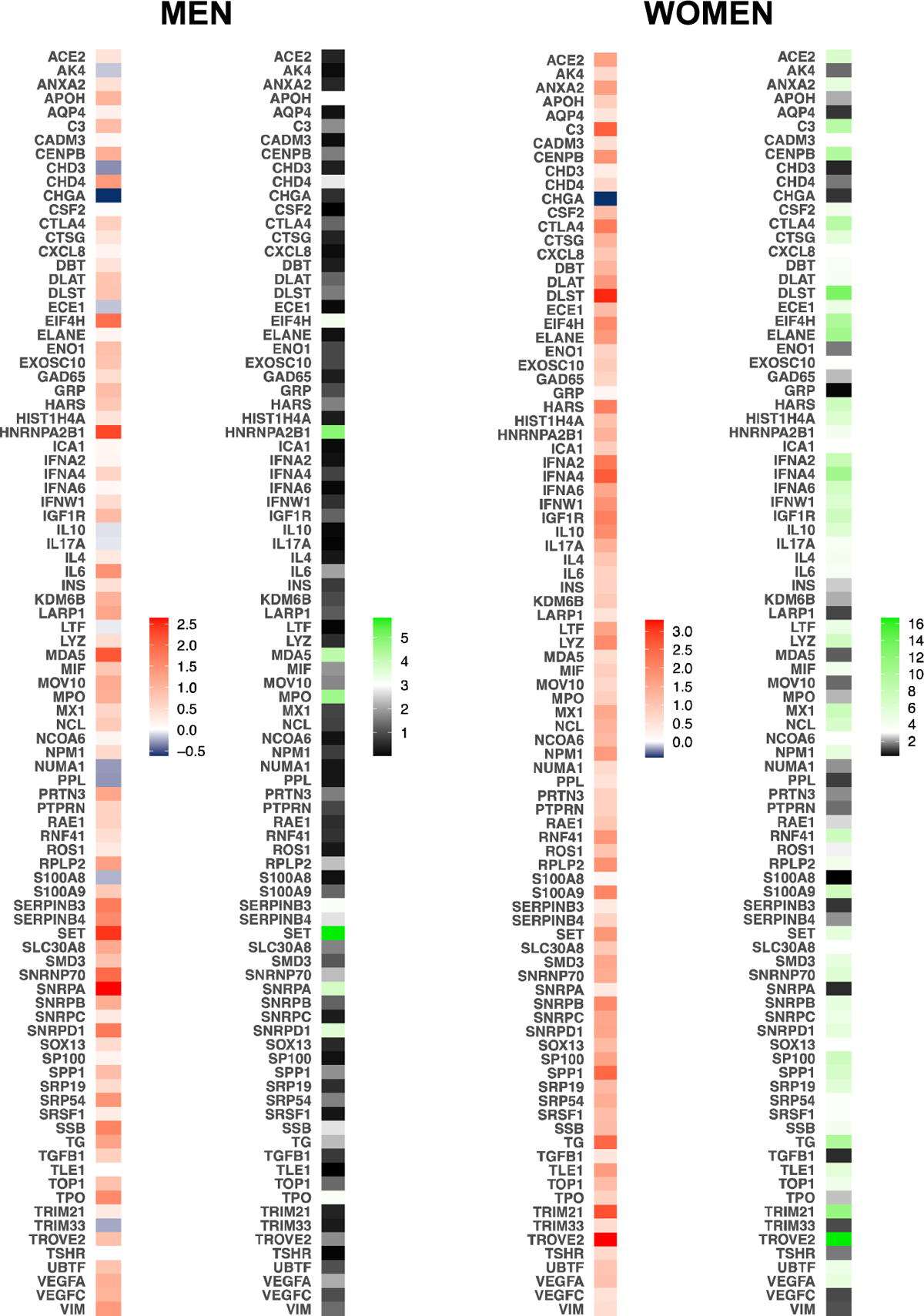
Sex-specific associations of autoantibodies with SLE status. In age-adjusted regression analyses, the breadth and magnitude of associations observed AABs reactivity and systemic lupus erythematosus (SLE) compared to health control status were predominantly seen in women compared to men. Beta coefficients were shown to the left, and negative log P values were shown to the right in each panel.

**Table S1.**
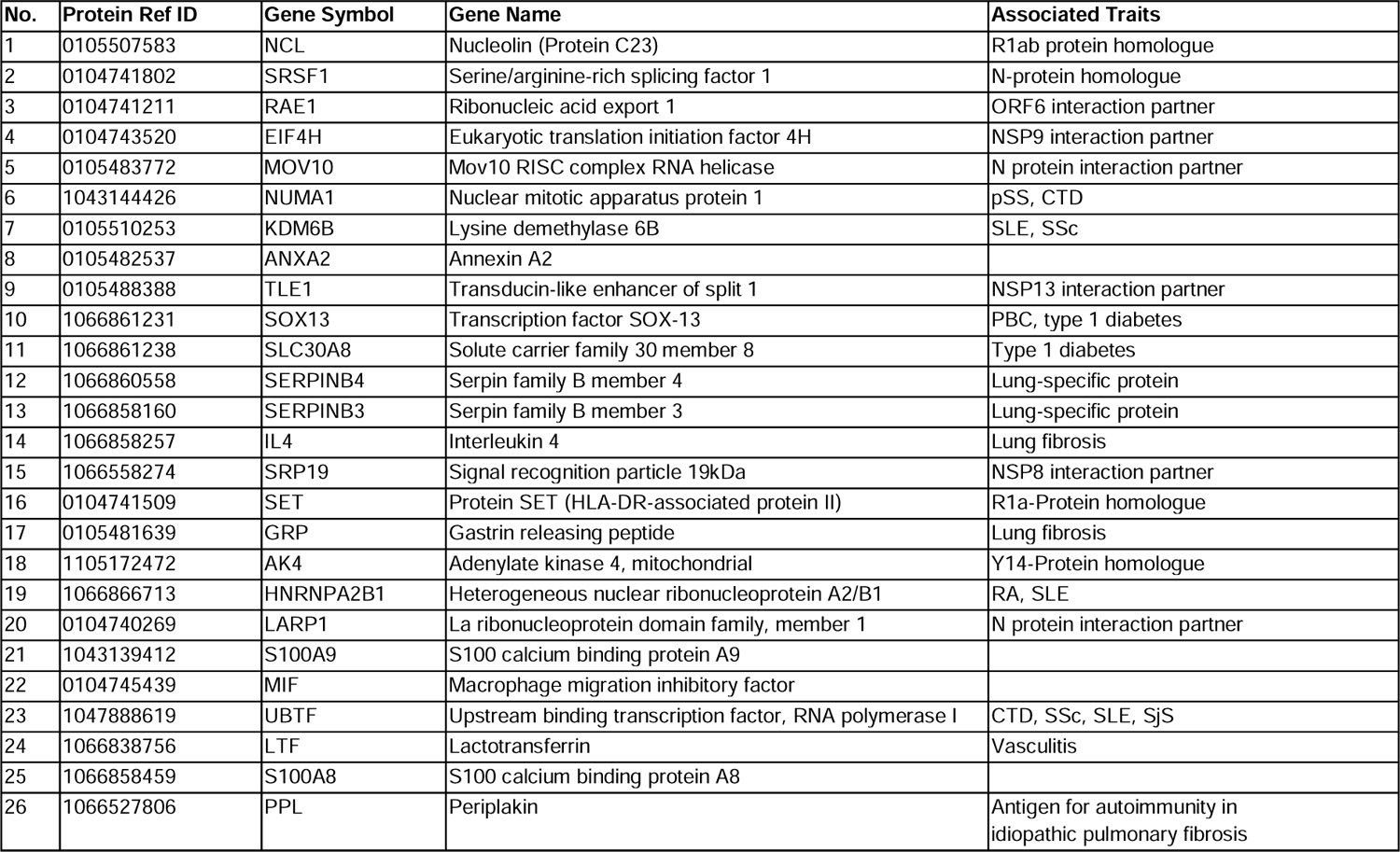
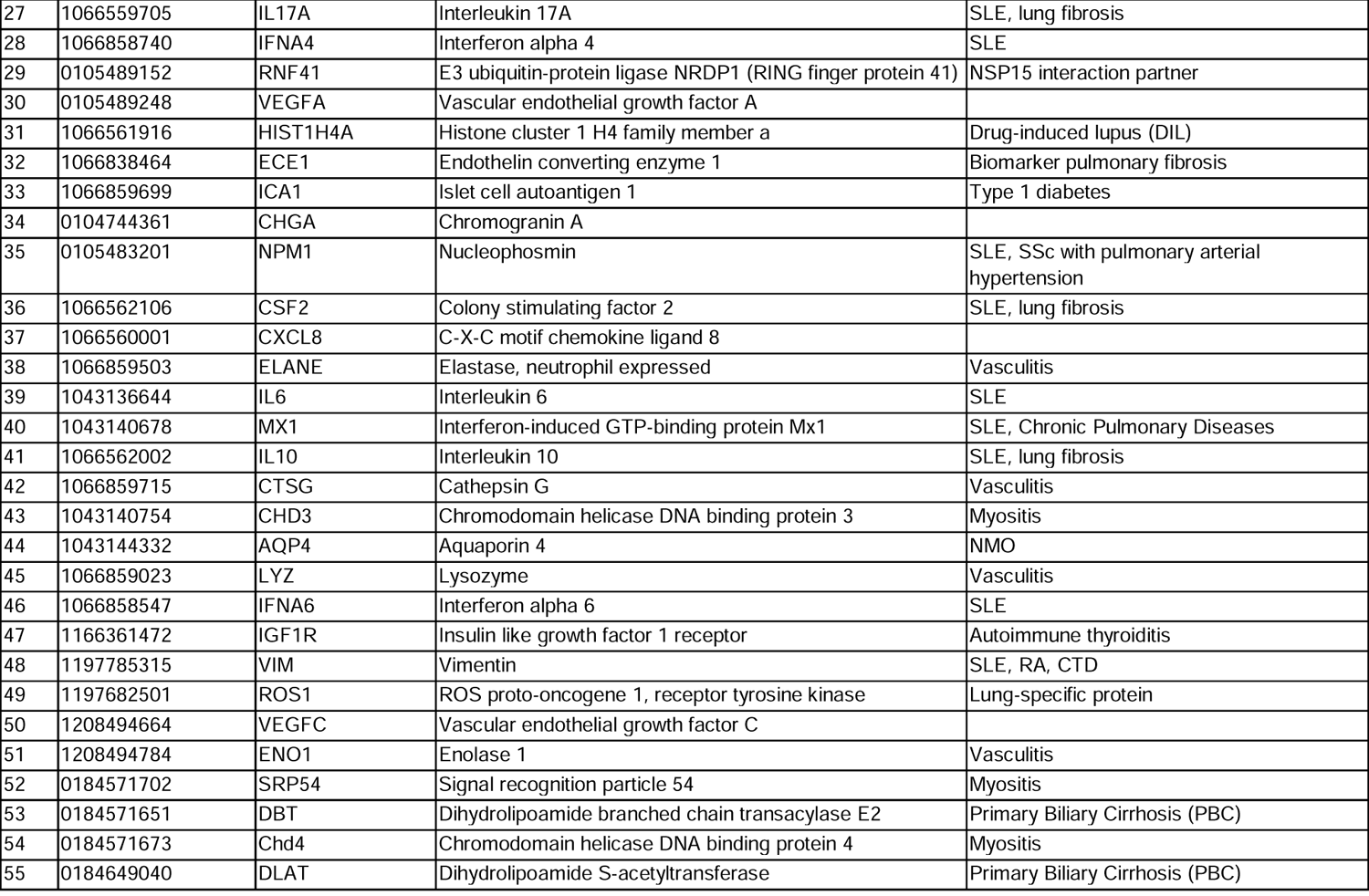
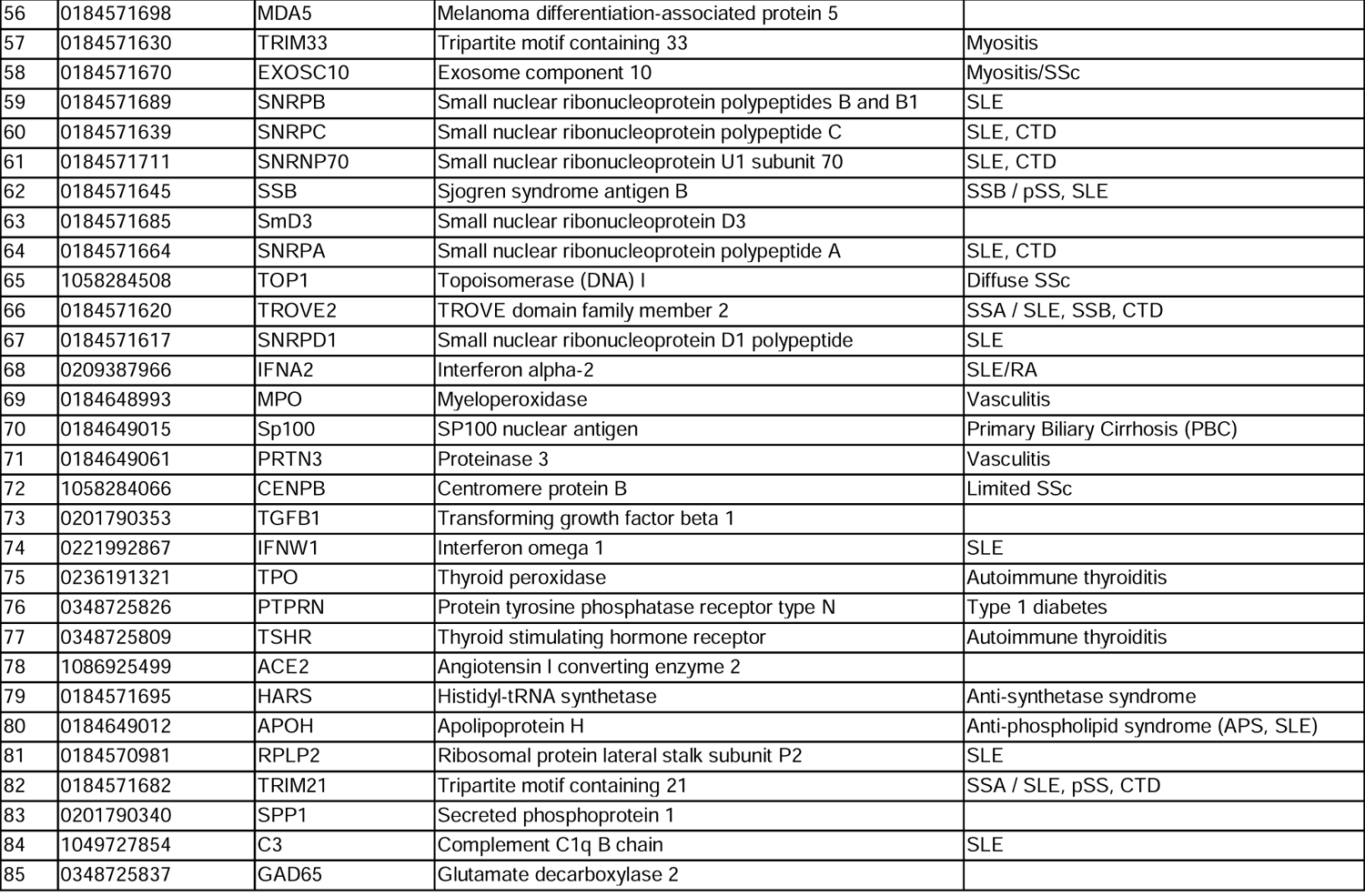
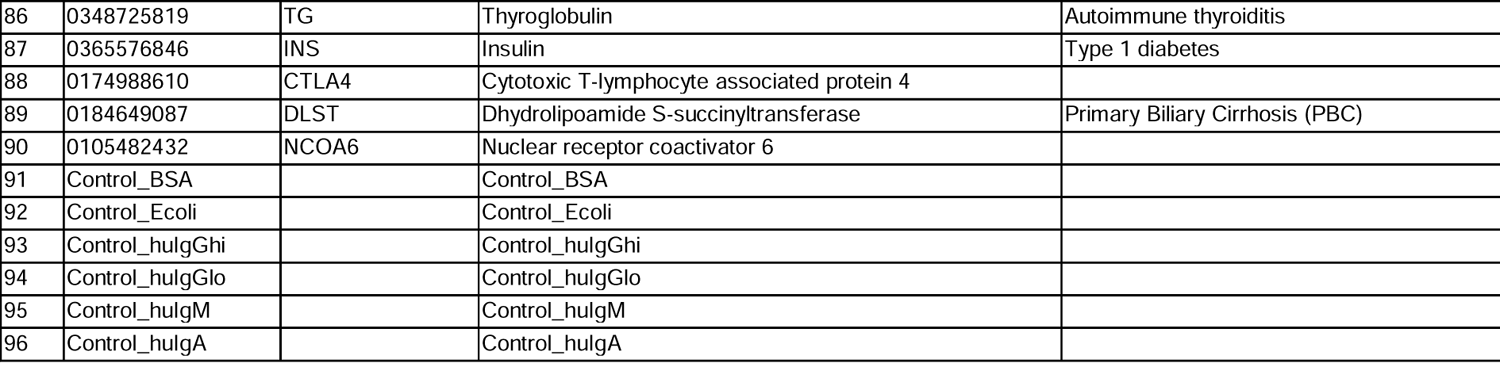
Composition of the AABs array.

**Table S2.**
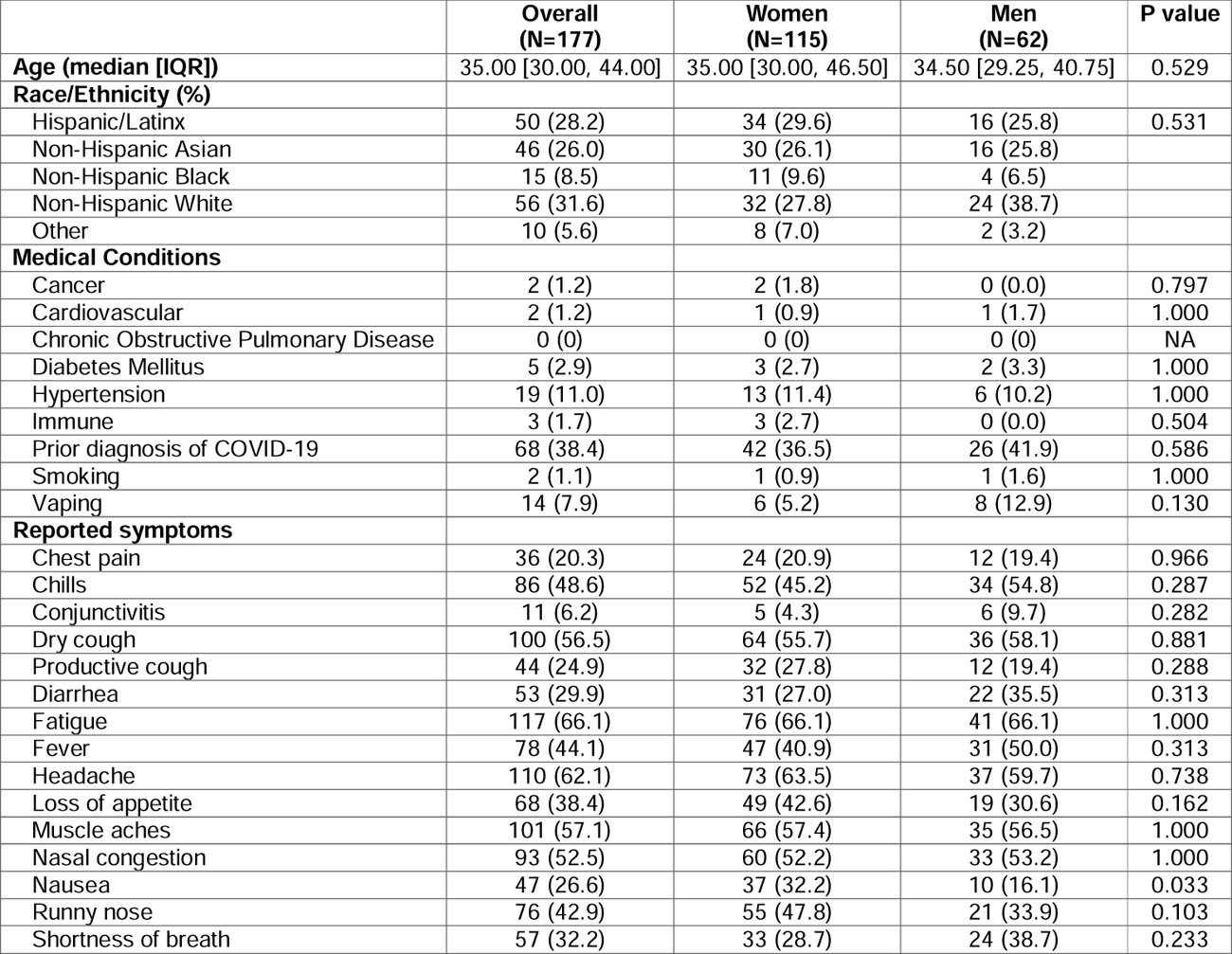
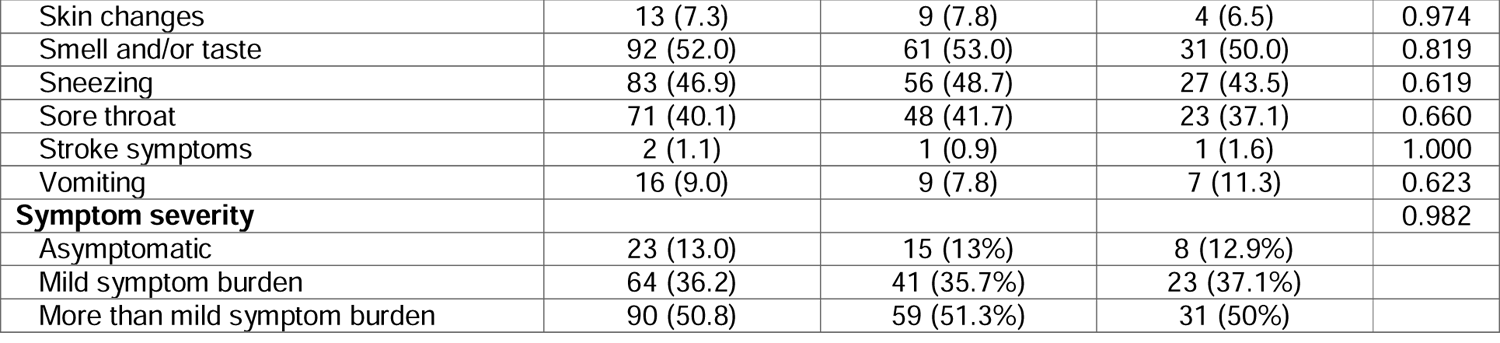
Demographic and clinical characteristics of 177 HCWs.

**Table S3.**
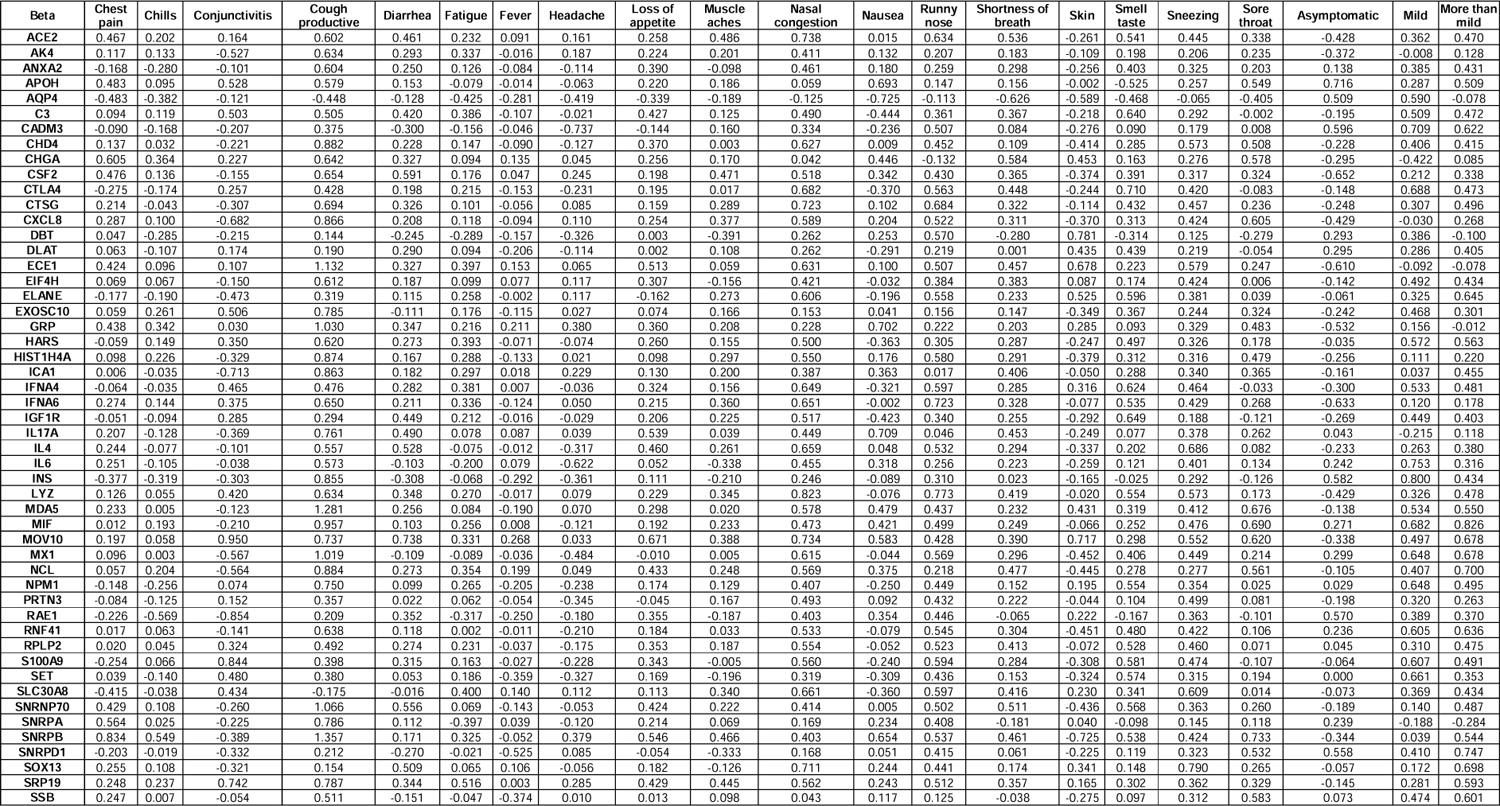
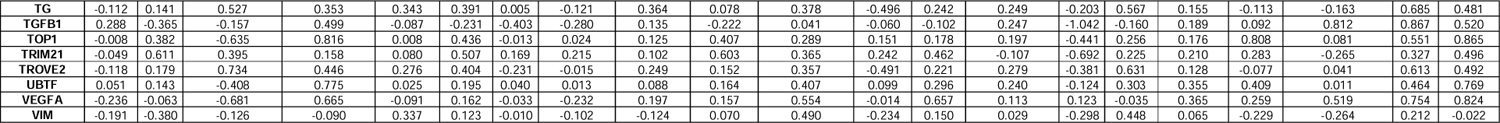
Associations of AABs reactivity with symptoms in men. Beta coefficients from age-adjusted regression analysis comparing males with a specific symptom burden to males without the same symptom are shown. Last three columns show beta coefficients from age-adjusted regression analysis comparing males with different levels of symptoms burdens to the pre-pandemic healthy control group.

**Table S4.**
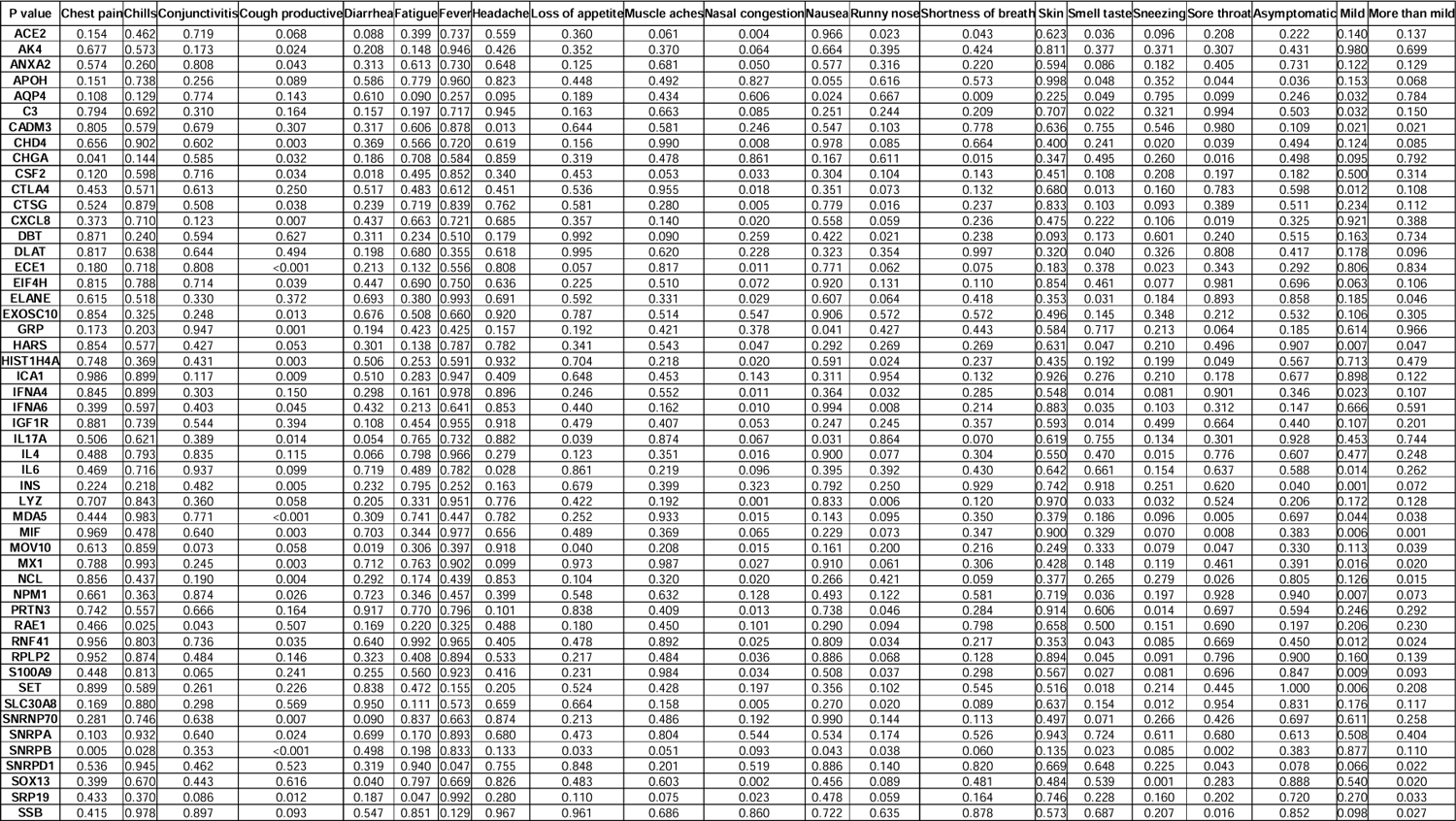
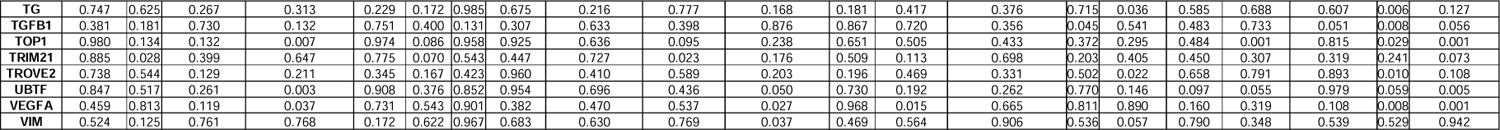
Associations of AABs reactivity with symptoms in men. P values from age-adjusted regression analysis comparing males with a specific symptom burden to males without the same symptom are shown. Last three columns show p values from age-adjusted regression analysis comparing males with different levels of symptoms burdens to the pre-pandemic healthy control group.

**Table S5.**
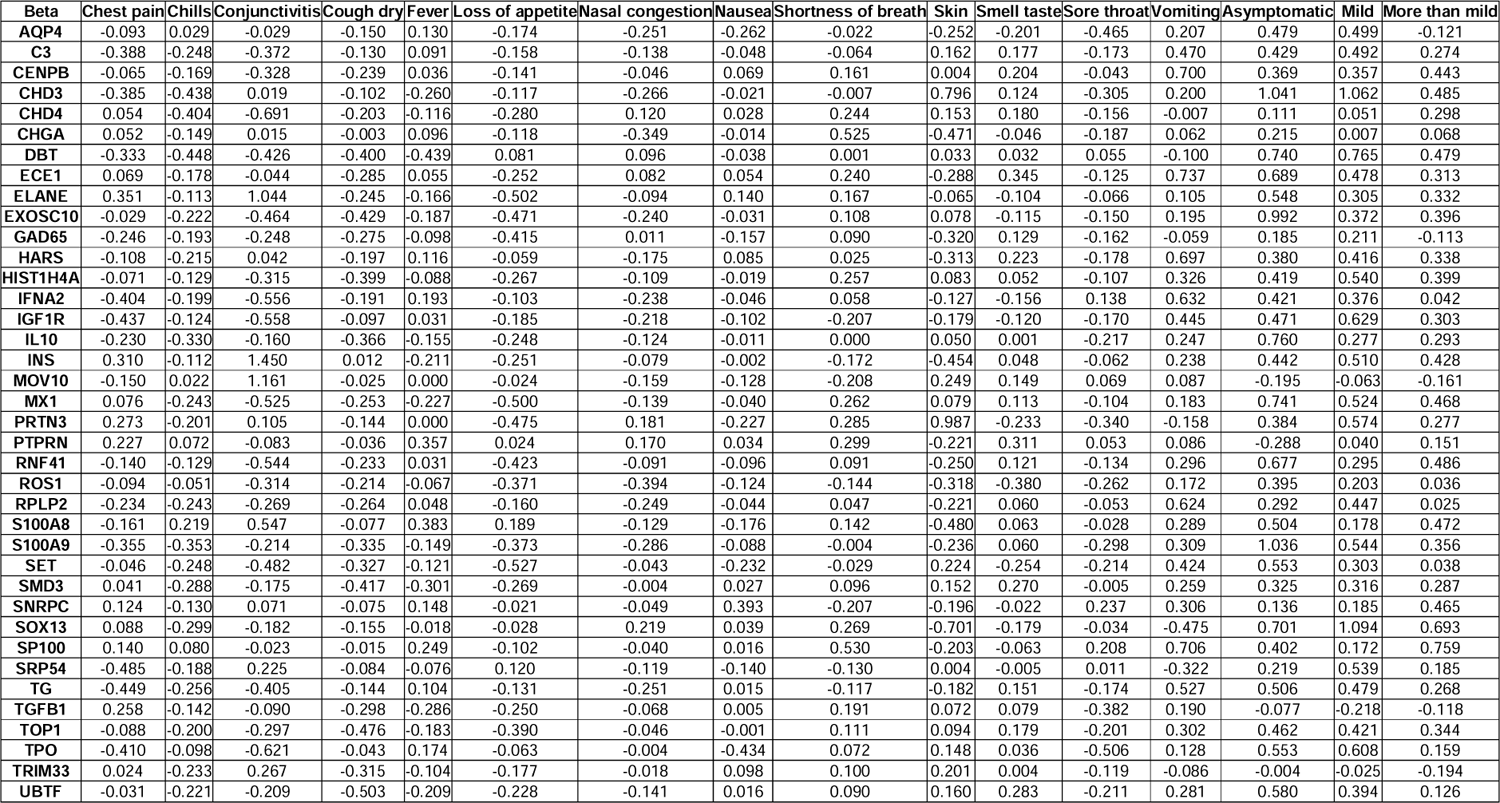
Associations of AABs reactivity with symptoms in women. Beta coefficients from age-adjusted regression analysis comparing males with a specific symptom burden to females without the same symptom are shown. Last three columns show beta coefficients from age-adjusted regression analysis comparing females with different levels of symptoms burdens to the pre-pandemic healthy control group.

**Table S6.**
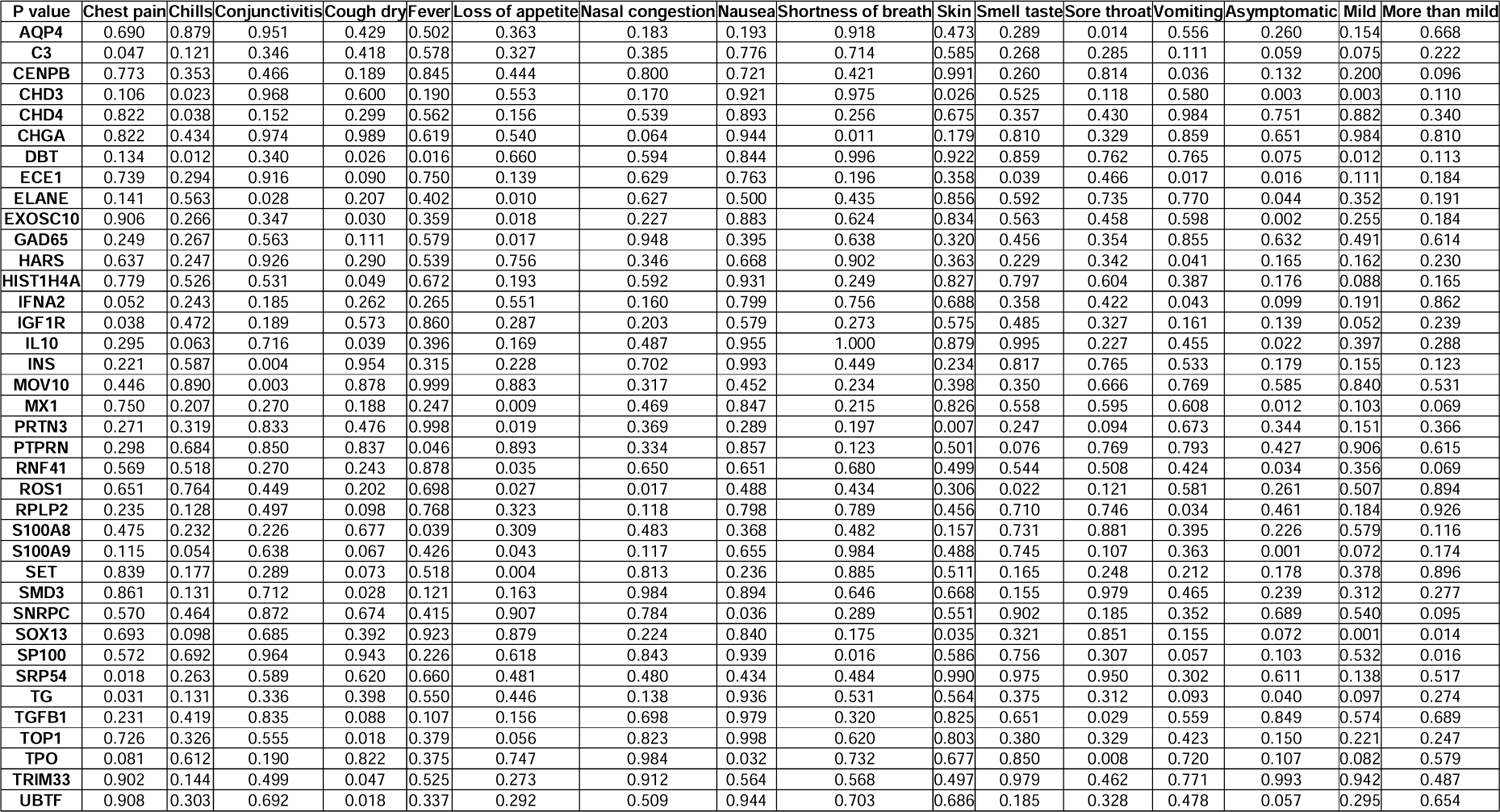
Associations of AABs reactivity with symptoms in women. P values from age-adjusted regression analysis comparing males with a specific symptom burden to females without the same symptom are shown. Last three columns show p values from age-adjusted regression analysis comparing females with different levels of symptoms burdens to the pre-pandemic healthy control group.

