## Supplemental Material for "Paradoxical Sex-Specific Patterns of Autoantibodies Response to SARS-CoV-2 Infection"

**Table S1.** Composition of the AABs array.

| **No.** | **Protein Ref ID** | **Gene Symbol** | **Gene Name** | **Associated Traits** |
| --- | --- | --- | --- | --- |
| 1 | 0105507583 | NCL | Nucleolin (Protein C23) | R1ab protein homologue |
| 2 | 0104741802 | SRSF1 | Serine/arginine-rich splicing factor 1 | N-protein homologue |
| 3 | 0104741211 | RAE1 | Ribonucleic acid export 1 | ORF6 interaction partner |
| 4 | 0104743520 | EIF4H | Eukaryotic translation initiation factor 4H | NSP9 interaction partner |
| 5 | 0105483772 | MOV10 | Mov10 RISC complex RNA helicase | N protein interaction partner |
| 6 | 1043144426 | NUMA1 | Nuclear mitotic apparatus protein 1 | pSS, CTD |
| 7 | 0105510253 | KDM6B | Lysine demethylase 6B | SLE, SSc |
| 8 | 0105482537 | ANXA2 | Annexin A2 |  |
| 9 | 0105488388 | TLE1 | Transducin-like enhancer of split 1 | NSP13 interaction partner |
| 10 | 1066861231 | SOX13 | Transcription factor SOX-13 | PBC, type 1 diabetes |
| 11 | 1066861238 | SLC30A8 | Solute carrier family 30 member 8 | Type 1 diabetes |
| 12 | 1066860558 | SERPINB4 | Serpin family B member 4 | Lung-specific protein |
| 13 | 1066858160 | SERPINB3 | Serpin family B member 3 | Lung-specific protein |
| 14 | 1066858257 | IL4 | Interleukin 4 | Lung fibrosis |
| 15 | 1066558274 | SRP19 | Signal recognition particle 19kDa | NSP8 interaction partner |
| 16 | 0104741509 | SET | Protein SET (HLA-DR-associated protein II) | R1a-Protein homologue |
| 17 | 0105481639 | GRP | Gastrin releasing peptide | Lung fibrosis |
| 18 | 1105172472 | AK4 | Adenylate kinase 4, mitochondrial | Y14-Protein homologue |
| 19 | 1066866713 | HNRNPA2B1 | Heterogeneous nuclear ribonucleoprotein A2/B1 | RA, SLE |
| 20 | 0104740269 | LARP1 | La ribonucleoprotein domain family, member 1 | N protein interaction partner |
| 21 | 1043139412 | S100A9 | S100 calcium binding protein A9 |  |
| 22 | 0104745439 | MIF | Macrophage migration inhibitory factor |  |
| 23 | 1047888619 | UBTF | Upstream binding transcription factor, RNA polymerase I | CTD, SSc, SLE, SjS |
| 24 | 1066838756 | LTF | Lactotransferrin | Vasculitis |
| 25 | 1066858459 | S100A8 | S100 calcium binding protein A8 |  |
| 26 | 1066527806 | PPL | Periplakin | Antigen for autoimmunity in idiopathic pulmonary fibrosis |
| 27 | 1066559705 | IL17A | Interleukin 17A | SLE, lung fibrosis |
| 28 | 1066858740 | IFNA4 | Interferon alpha 4 | SLE |
| 29 | 0105489152 | RNF41 | E3 ubiquitin-protein ligase NRDP1 (RING finger protein 41) | NSP15 interaction partner |
| 30 | 0105489248 | VEGFA | Vascular endothelial growth factor A |  |
| 31 | 1066561916 | HIST1H4A | Histone cluster 1 H4 family member a | Drug-induced lupus (DIL) |
| 32 | 1066838464 | ECE1 | Endothelin converting enzyme 1 | Biomarker pulmonary fibrosis |
| 33 | 1066859699 | ICA1 | Islet cell autoantigen 1 | Type 1 diabetes |
| 34 | 0104744361 | CHGA | Chromogranin A |  |
| 35 | 0105483201 | NPM1 | Nucleophosmin | SLE, SSc with pulmonary arterial hypertension |
| 36 | 1066562106 | CSF2 | Colony stimulating factor 2 | SLE, lung fibrosis |
| 37 | 1066560001 | CXCL8 | C-X-C motif chemokine ligand 8 |  |
| 38 | 1066859503 | ELANE | Elastase, neutrophil expressed | Vasculitis |
| 39 | 1043136644 | IL6 | Interleukin 6 | SLE |
| 40 | 1043140678 | MX1 | Interferon-induced GTP-binding protein Mx1 | SLE, Chronic Pulmonary Diseases |
| 41 | 1066562002 | IL10 | Interleukin 10 | SLE, lung fibrosis |
| 42 | 1066859715 | CTSG | Cathepsin G | Vasculitis |
| 43 | 1043140754 | CHD3 | Chromodomain helicase DNA binding protein 3 | Myositis |
| 44 | 1043144332 | AQP4 | Aquaporin 4 | NMO |
| 45 | 1066859023 | LYZ | Lysozyme | Vasculitis |
| 46 | 1066858547 | IFNA6 | Interferon alpha 6 | SLE |
| 47 | 1166361472 | IGF1R | Insulin like growth factor 1 receptor | Autoimmune thyroiditis |
| 48 | 1197785315 | VIM | Vimentin | SLE, RA, CTD |
| 49 | 1197682501 | ROS1 | ROS proto-oncogene 1, receptor tyrosine kinase | Lung-specific protein |
| 50 | 1208494664 | VEGFC | Vascular endothelial growth factor C |  |
| 51 | 1208494784 | ENO1 | Enolase 1 | Vasculitis |
| 52 | 0184571702 | SRP54 | Signal recognition particle 54 | Myositis |
| 53 | 0184571651 | DBT | Dihydrolipoamide branched chain transacylase E2 | Primary Biliary Cirrhosis (PBC) |
| 54 | 0184571673 | Chd4 | Chromodomain helicase DNA binding protein 4 | Myositis |
| 55 | 0184649040 | DLAT | Dihydrolipoamide S-acetyltransferase | Primary Biliary Cirrhosis (PBC) |
| 56 | 0184571698 | MDA5 | Melanoma differentiation-associated protein 5 |  |
| 57 | 0184571630 | TRIM33 | Tripartite motif containing 33 | Myositis |
| 58 | 0184571670 | EXOSC10 | Exosome component 10 | Myositis/SSc |
| 59 | 0184571689 | SNRPB | Small nuclear ribonucleoprotein polypeptides B and B1 | SLE |
| 60 | 0184571639 | SNRPC | Small nuclear ribonucleoprotein polypeptide C | SLE, CTD |
| 61 | 0184571711 | SNRNP70 | Small nuclear ribonucleoprotein U1 subunit 70 | SLE, CTD |
| 62 | 0184571645 | SSB | Sjogren syndrome antigen B | SSB / pSS, SLE |
| 63 | 0184571685 | SmD3 | Small nuclear ribonucleoprotein D3 |  |
| 64 | 0184571664 | SNRPA | Small nuclear ribonucleoprotein polypeptide A | SLE, CTD |
| 65 | 1058284508 | TOP1 | Topoisomerase (DNA) I | Diffuse SSc |
| 66 | 0184571620 | TROVE2 | TROVE domain family member 2 | SSA / SLE, SSB, CTD |
| 67 | 0184571617 | SNRPD1 | Small nuclear ribonucleoprotein D1 polypeptide | SLE |
| 68 | 0209387966 | IFNA2 | Interferon alpha-2 | SLE/RA |
| 69 | 0184648993 | MPO | Myeloperoxidase | Vasculitis |
| 70 | 0184649015 | Sp100 | SP100 nuclear antigen | Primary Biliary Cirrhosis (PBC) |
| 71 | 0184649061 | PRTN3 | Proteinase 3 | Vasculitis |
| 72 | 1058284066 | CENPB | Centromere protein B | Limited SSc |
| 73 | 0201790353 | TGFB1 | Transforming growth factor beta 1 |  |
| 74 | 0221992867 | IFNW1 | Interferon omega 1 | SLE |
| 75 | 0236191321 | TPO | Thyroid peroxidase | Autoimmune thyroiditis |
| 76 | 0348725826 | PTPRN | Protein tyrosine phosphatase receptor type N | Type 1 diabetes |
| 77 | 0348725809 | TSHR | Thyroid stimulating hormone receptor | Autoimmune thyroiditis |
| 78 | 1086925499 | ACE2 | Angiotensin I converting enzyme 2 |  |
| 79 | 0184571695 | HARS | Histidyl-tRNA synthetase | Anti-synthetase syndrome |
| 80 | 0184649012 | APOH | Apolipoprotein H | Anti-phospholipid syndrome (APS, SLE) |
| 81 | 0184570981 | RPLP2 | Ribosomal protein lateral stalk subunit P2 | SLE |
| 82 | 0184571682 | TRIM21 | Tripartite motif containing 21 | SSA / SLE, pSS, CTD |
| 83 | 0201790340 | SPP1 | Secreted phosphoprotein 1 |  |
| 84 | 1049727854 | C3 | Complement C1q B chain | SLE |
| 85 | 0348725837 | GAD65 | Glutamate decarboxylase 2 |  |
| 86 | 0348725819 | TG | Thyroglobulin | Autoimmune thyroiditis |
| 87 | 0365576846 | INS | Insulin | Type 1 diabetes |
| 88 | 0174988610 | CTLA4 | Cytotoxic T-lymphocyte associated protein 4 |  |
| 89 | 0184649087 | DLST | Dhydrolipoamide S-succinyltransferase | Primary Biliary Cirrhosis (PBC) |
| 90 | 0105482432 | NCOA6 | Nuclear receptor coactivator 6 |  |
| 91 | Control_BSA |  | Control_BSA |  |
| 92 | Control_Ecoli |  | Control_Ecoli |  |
| 93 | Control_huIgGhi |  | Control_huIgGhi |  |
| 94 | Control_huIgGlo |  | Control_huIgGlo |  |
| 95 | Control_huIgM |  | Control_huIgM |  |
| 96 | Control_huIgA |  | Control_huIgA |  |

**Table S2.** Demographic and clinical characteristics of 177 HCWs.

|  | **Overall**  **(N=177)** | **Women**  **(N=115)** | **Men**  **(N=62)** | **P value** |
| --- | --- | --- | --- | --- |
| **Age (median [IQR])** | 35.00 [30.00, 44.00] | 35.00 [30.00, 46.50] | 34.50 [29.25, 40.75] | 0.529 |
| **Race/Ethnicity (%)** |  |  |  |  |
| Hispanic/Latinx | 50 (28.2) | 34 (29.6) | 16 (25.8) | 0.531 |
| Non-Hispanic Asian | 46 (26.0) | 30 (26.1) | 16 (25.8) |  |
| Non-Hispanic Black | 15 (8.5) | 11 (9.6) | 4 (6.5) |  |
| Non-Hispanic White | 56 (31.6) | 32 (27.8) | 24 (38.7) |  |
| Other | 10 (5.6) | 8 (7.0) | 2 (3.2) |  |
| **Medical Conditions** |  |  |  |  |
| Cancer | 2 (1.2) | 2 (1.8) | 0 (0.0) | 0.797 |
| Cardiovascular | 2 (1.2) | 1 (0.9) | 1 (1.7) | 1.000 |
| Chronic Obstructive Pulmonary Disease | 0 (0) | 0 (0) | 0 (0) | NA |
| Diabetes Mellitus | 5 (2.9) | 3 (2.7) | 2 (3.3) | 1.000 |
| Hypertension | 19 (11.0) | 13 (11.4) | 6 (10.2) | 1.000 |
| Immune | 3 (1.7) | 3 (2.7) | 0 (0.0) | 0.504 |
| Prior diagnosis of COVID-19 | 68 (38.4) | 42 (36.5) | 26 (41.9) | 0.586 |
| Smoking | 2 (1.1) | 1 (0.9) | 1 (1.6) | 1.000 |
| Vaping | 14 (7.9) | 6 (5.2) | 8 (12.9) | 0.130 |
| **Reported symptoms** |  |  |  |  |
| Chest pain | 36 (20.3) | 24 (20.9) | 12 (19.4) | 0.966 |
| Chills | 86 (48.6) | 52 (45.2) | 34 (54.8) | 0.287 |
| Conjunctivitis | 11 (6.2) | 5 (4.3) | 6 (9.7) | 0.282 |
| Dry cough | 100 (56.5) | 64 (55.7) | 36 (58.1) | 0.881 |
| Productive cough | 44 (24.9) | 32 (27.8) | 12 (19.4) | 0.288 |
| Diarrhea | 53 (29.9) | 31 (27.0) | 22 (35.5) | 0.313 |
| Fatigue | 117 (66.1) | 76 (66.1) | 41 (66.1) | 1.000 |
| Fever | 78 (44.1) | 47 (40.9) | 31 (50.0) | 0.313 |
| Headache | 110 (62.1) | 73 (63.5) | 37 (59.7) | 0.738 |
| Loss of appetite | 68 (38.4) | 49 (42.6) | 19 (30.6) | 0.162 |
| Muscle aches | 101 (57.1) | 66 (57.4) | 35 (56.5) | 1.000 |
| Nasal congestion | 93 (52.5) | 60 (52.2) | 33 (53.2) | 1.000 |
| Nausea | 47 (26.6) | 37 (32.2) | 10 (16.1) | 0.033 |
| Runny nose | 76 (42.9) | 55 (47.8) | 21 (33.9) | 0.103 |
| Shortness of breath | 57 (32.2) | 33 (28.7) | 24 (38.7) | 0.233 |
| Skin changes | 13 (7.3) | 9 (7.8) | 4 (6.5) | 0.974 |
| Smell and/or taste | 92 (52.0) | 61 (53.0) | 31 (50.0) | 0.819 |
| Sneezing | 83 (46.9) | 56 (48.7) | 27 (43.5) | 0.619 |
| Sore throat | 71 (40.1) | 48 (41.7) | 23 (37.1) | 0.660 |
| Stroke symptoms | 2 (1.1) | 1 (0.9) | 1 (1.6) | 1.000 |
| Vomiting | 16 (9.0) | 9 (7.8) | 7 (11.3) | 0.623 |
| **Symptom severity** |  |  |  | 0.982 |
| Asymptomatic | 23 (13.0) | 15 (13%) | 8 (12.9%) |  |
| Mild symptom burden | 64 (36.2) | 41 (35.7%) | 23 (37.1%) |  |
| More than mild symptom burden | 90 (50.8) | 59 (51.3%) | 31 (50%) |  |

**Table S3.** Associations of AABs reactivity with symptoms in men. Beta coefficients from age-adjusted regression analysis comparing males with a specific symptom burden to males without the same symptom are shown. Last three columns show beta coefficients from age-adjusted regression analysis comparing males with different levels of symptoms burdens to the pre-pandemic healthy control group.

| **Beta** | **Chest pain** | **Chills** | **Conjunctivitis** | **Cough productive** | **Diarrhea** | **Fatigue** | **Fever** | **Headache** | **Loss of appetite** | **Muscle aches** | **Nasal congestion** | **Nausea** | **Runny nose** | **Shortness of breath** | **Skin** | **Smell taste** | **Sneezing** | **Sore throat** | **Asymptomatic** | **Mild** | **More than mild** |
| --- | --- | --- | --- | --- | --- | --- | --- | --- | --- | --- | --- | --- | --- | --- | --- | --- | --- | --- | --- | --- | --- |
| **ACE2** | 0.467 | 0.202 | 0.164 | 0.602 | 0.461 | 0.232 | 0.091 | 0.161 | 0.258 | 0.486 | 0.738 | 0.015 | 0.634 | 0.536 | -0.261 | 0.541 | 0.445 | 0.338 | -0.428 | 0.362 | 0.470 |
| **AK4** | 0.117 | 0.133 | -0.527 | 0.634 | 0.293 | 0.337 | -0.016 | 0.187 | 0.224 | 0.201 | 0.411 | 0.132 | 0.207 | 0.183 | -0.109 | 0.198 | 0.206 | 0.235 | -0.372 | -0.008 | 0.128 |
| **ANXA2** | -0.168 | -0.280 | -0.101 | 0.604 | 0.250 | 0.126 | -0.084 | -0.114 | 0.390 | -0.098 | 0.461 | 0.180 | 0.259 | 0.298 | -0.256 | 0.403 | 0.325 | 0.203 | 0.138 | 0.385 | 0.431 |
| **APOH** | 0.483 | 0.095 | 0.528 | 0.579 | 0.153 | -0.079 | -0.014 | -0.063 | 0.220 | 0.186 | 0.059 | 0.693 | 0.147 | 0.156 | -0.002 | -0.525 | 0.257 | 0.549 | 0.716 | 0.287 | 0.509 |
| **AQP4** | -0.483 | -0.382 | -0.121 | -0.448 | -0.128 | -0.425 | -0.281 | -0.419 | -0.339 | -0.189 | -0.125 | -0.725 | -0.113 | -0.626 | -0.589 | -0.468 | -0.065 | -0.405 | 0.509 | 0.590 | -0.078 |
| **C3** | 0.094 | 0.119 | 0.503 | 0.505 | 0.420 | 0.386 | -0.107 | -0.021 | 0.427 | 0.125 | 0.490 | -0.444 | 0.361 | 0.367 | -0.218 | 0.640 | 0.292 | -0.002 | -0.195 | 0.509 | 0.472 |
| **CADM3** | -0.090 | -0.168 | -0.207 | 0.375 | -0.300 | -0.156 | -0.046 | -0.737 | -0.144 | 0.160 | 0.334 | -0.236 | 0.507 | 0.084 | -0.276 | 0.090 | 0.179 | 0.008 | 0.596 | 0.709 | 0.622 |
| **CHD4** | 0.137 | 0.032 | -0.221 | 0.882 | 0.228 | 0.147 | -0.090 | -0.127 | 0.370 | 0.003 | 0.627 | 0.009 | 0.452 | 0.109 | -0.414 | 0.285 | 0.573 | 0.508 | -0.228 | 0.406 | 0.415 |
| **CHGA** | 0.605 | 0.364 | 0.227 | 0.642 | 0.327 | 0.094 | 0.135 | 0.045 | 0.256 | 0.170 | 0.042 | 0.446 | -0.132 | 0.584 | 0.453 | 0.163 | 0.276 | 0.578 | -0.295 | -0.422 | 0.085 |
| **CSF2** | 0.476 | 0.136 | -0.155 | 0.654 | 0.591 | 0.176 | 0.047 | 0.245 | 0.198 | 0.471 | 0.518 | 0.342 | 0.430 | 0.365 | -0.374 | 0.391 | 0.317 | 0.324 | -0.652 | 0.212 | 0.338 |
| **CTLA4** | -0.275 | -0.174 | 0.257 | 0.428 | 0.198 | 0.215 | -0.153 | -0.231 | 0.195 | 0.017 | 0.682 | -0.370 | 0.563 | 0.448 | -0.244 | 0.710 | 0.420 | -0.083 | -0.148 | 0.688 | 0.473 |
| **CTSG** | 0.214 | -0.043 | -0.307 | 0.694 | 0.326 | 0.101 | -0.056 | 0.085 | 0.159 | 0.289 | 0.723 | 0.102 | 0.684 | 0.322 | -0.114 | 0.432 | 0.457 | 0.236 | -0.248 | 0.307 | 0.496 |
| **CXCL8** | 0.287 | 0.100 | -0.682 | 0.866 | 0.208 | 0.118 | -0.094 | 0.110 | 0.254 | 0.377 | 0.589 | 0.204 | 0.522 | 0.311 | -0.370 | 0.313 | 0.424 | 0.605 | -0.429 | -0.030 | 0.268 |
| **DBT** | 0.047 | -0.285 | -0.215 | 0.144 | -0.245 | -0.289 | -0.157 | -0.326 | 0.003 | -0.391 | 0.262 | 0.253 | 0.570 | -0.280 | 0.781 | -0.314 | 0.125 | -0.279 | 0.293 | 0.386 | -0.100 |
| **DLAT** | 0.063 | -0.107 | 0.174 | 0.190 | 0.290 | 0.094 | -0.206 | -0.114 | 0.002 | 0.108 | 0.262 | -0.291 | 0.219 | 0.001 | 0.435 | 0.439 | 0.219 | -0.054 | 0.295 | 0.286 | 0.405 |
| **ECE1** | 0.424 | 0.096 | 0.107 | 1.132 | 0.327 | 0.397 | 0.153 | 0.065 | 0.513 | 0.059 | 0.631 | 0.100 | 0.507 | 0.457 | 0.678 | 0.223 | 0.579 | 0.247 | -0.610 | -0.092 | -0.078 |
| **EIF4H** | 0.069 | 0.067 | -0.150 | 0.612 | 0.187 | 0.099 | 0.077 | 0.117 | 0.307 | -0.156 | 0.421 | -0.032 | 0.384 | 0.383 | 0.087 | 0.174 | 0.424 | 0.006 | -0.142 | 0.492 | 0.434 |
| **ELANE** | -0.177 | -0.190 | -0.473 | 0.319 | 0.115 | 0.258 | -0.002 | 0.117 | -0.162 | 0.273 | 0.606 | -0.196 | 0.558 | 0.233 | 0.525 | 0.596 | 0.381 | 0.039 | -0.061 | 0.325 | 0.645 |
| **EXOSC10** | 0.059 | 0.261 | 0.506 | 0.785 | -0.111 | 0.176 | -0.115 | 0.027 | 0.074 | 0.166 | 0.153 | 0.041 | 0.156 | 0.147 | -0.349 | 0.367 | 0.244 | 0.324 | -0.242 | 0.468 | 0.301 |
| **GRP** | 0.438 | 0.342 | 0.030 | 1.030 | 0.347 | 0.216 | 0.211 | 0.380 | 0.360 | 0.208 | 0.228 | 0.702 | 0.222 | 0.203 | 0.285 | 0.093 | 0.329 | 0.483 | -0.532 | 0.156 | -0.012 |
| **HARS** | -0.059 | 0.149 | 0.350 | 0.620 | 0.273 | 0.393 | -0.071 | -0.074 | 0.260 | 0.155 | 0.500 | -0.363 | 0.305 | 0.287 | -0.247 | 0.497 | 0.326 | 0.178 | -0.035 | 0.572 | 0.563 |
| **HIST1H4A** | 0.098 | 0.226 | -0.329 | 0.874 | 0.167 | 0.288 | -0.133 | 0.021 | 0.098 | 0.297 | 0.550 | 0.176 | 0.580 | 0.291 | -0.379 | 0.312 | 0.316 | 0.479 | -0.256 | 0.111 | 0.220 |
| **ICA1** | 0.006 | -0.035 | -0.713 | 0.863 | 0.182 | 0.297 | 0.018 | 0.229 | 0.130 | 0.200 | 0.387 | 0.363 | 0.017 | 0.406 | -0.050 | 0.288 | 0.340 | 0.365 | -0.161 | 0.037 | 0.455 |
| **IFNA4** | -0.064 | -0.035 | 0.465 | 0.476 | 0.282 | 0.381 | 0.007 | -0.036 | 0.324 | 0.156 | 0.649 | -0.321 | 0.597 | 0.285 | 0.316 | 0.624 | 0.464 | -0.033 | -0.300 | 0.533 | 0.481 |
| **IFNA6** | 0.274 | 0.144 | 0.375 | 0.650 | 0.211 | 0.336 | -0.124 | 0.050 | 0.215 | 0.360 | 0.651 | -0.002 | 0.723 | 0.328 | -0.077 | 0.535 | 0.429 | 0.268 | -0.633 | 0.120 | 0.178 |
| **IGF1R** | -0.051 | -0.094 | 0.285 | 0.294 | 0.449 | 0.212 | -0.016 | -0.029 | 0.206 | 0.225 | 0.517 | -0.423 | 0.340 | 0.255 | -0.292 | 0.649 | 0.188 | -0.121 | -0.269 | 0.449 | 0.403 |
| **IL17A** | 0.207 | -0.128 | -0.369 | 0.761 | 0.490 | 0.078 | 0.087 | 0.039 | 0.539 | 0.039 | 0.449 | 0.709 | 0.046 | 0.453 | -0.249 | 0.077 | 0.378 | 0.262 | 0.043 | -0.215 | 0.118 |
| **IL4** | 0.244 | -0.077 | -0.101 | 0.557 | 0.528 | -0.075 | -0.012 | -0.317 | 0.460 | 0.261 | 0.659 | 0.048 | 0.532 | 0.294 | -0.337 | 0.202 | 0.686 | 0.082 | -0.233 | 0.263 | 0.380 |
| **IL6** | 0.251 | -0.105 | -0.038 | 0.573 | -0.103 | -0.200 | 0.079 | -0.622 | 0.052 | -0.338 | 0.455 | 0.318 | 0.256 | 0.223 | -0.259 | 0.121 | 0.401 | 0.134 | 0.242 | 0.753 | 0.316 |
| **INS** | -0.377 | -0.319 | -0.303 | 0.855 | -0.308 | -0.068 | -0.292 | -0.361 | 0.111 | -0.210 | 0.246 | -0.089 | 0.310 | 0.023 | -0.165 | -0.025 | 0.292 | -0.126 | 0.582 | 0.800 | 0.434 |
| **LYZ** | 0.126 | 0.055 | 0.420 | 0.634 | 0.348 | 0.270 | -0.017 | 0.079 | 0.229 | 0.345 | 0.823 | -0.076 | 0.773 | 0.419 | -0.020 | 0.554 | 0.573 | 0.173 | -0.429 | 0.326 | 0.478 |
| **MDA5** | 0.233 | 0.005 | -0.123 | 1.281 | 0.256 | 0.084 | -0.190 | 0.070 | 0.298 | 0.020 | 0.578 | 0.479 | 0.437 | 0.232 | 0.431 | 0.319 | 0.412 | 0.676 | -0.138 | 0.534 | 0.550 |
| **MIF** | 0.012 | 0.193 | -0.210 | 0.957 | 0.103 | 0.256 | 0.008 | -0.121 | 0.192 | 0.233 | 0.473 | 0.421 | 0.499 | 0.249 | -0.066 | 0.252 | 0.476 | 0.690 | 0.271 | 0.682 | 0.826 |
| **MOV10** | 0.197 | 0.058 | 0.950 | 0.737 | 0.738 | 0.331 | 0.268 | 0.033 | 0.671 | 0.388 | 0.734 | 0.583 | 0.428 | 0.390 | 0.717 | 0.298 | 0.552 | 0.620 | -0.338 | 0.497 | 0.678 |
| **MX1** | 0.096 | 0.003 | -0.567 | 1.019 | -0.109 | -0.089 | -0.036 | -0.484 | -0.010 | 0.005 | 0.615 | -0.044 | 0.569 | 0.296 | -0.452 | 0.406 | 0.449 | 0.214 | 0.299 | 0.648 | 0.678 |
| **NCL** | 0.057 | 0.204 | -0.564 | 0.884 | 0.273 | 0.354 | 0.199 | 0.049 | 0.433 | 0.248 | 0.569 | 0.375 | 0.218 | 0.477 | -0.445 | 0.278 | 0.277 | 0.561 | -0.105 | 0.407 | 0.700 |
| **NPM1** | -0.148 | -0.256 | 0.074 | 0.750 | 0.099 | 0.265 | -0.205 | -0.238 | 0.174 | 0.129 | 0.407 | -0.250 | 0.449 | 0.152 | 0.195 | 0.554 | 0.354 | 0.025 | 0.029 | 0.648 | 0.495 |
| **PRTN3** | -0.084 | -0.125 | 0.152 | 0.357 | 0.022 | 0.062 | -0.054 | -0.345 | -0.045 | 0.167 | 0.493 | 0.092 | 0.432 | 0.222 | -0.044 | 0.104 | 0.499 | 0.081 | -0.198 | 0.320 | 0.263 |
| **RAE1** | -0.226 | -0.569 | -0.854 | 0.209 | 0.352 | -0.317 | -0.250 | -0.180 | 0.355 | -0.187 | 0.403 | 0.354 | 0.446 | -0.065 | 0.222 | -0.167 | 0.363 | -0.101 | 0.570 | 0.389 | 0.370 |
| **RNF41** | 0.017 | 0.063 | -0.141 | 0.638 | 0.118 | 0.002 | -0.011 | -0.210 | 0.184 | 0.033 | 0.533 | -0.079 | 0.545 | 0.304 | -0.451 | 0.480 | 0.422 | 0.106 | 0.236 | 0.605 | 0.636 |
| **RPLP2** | 0.020 | 0.045 | 0.324 | 0.492 | 0.274 | 0.231 | -0.037 | -0.175 | 0.353 | 0.187 | 0.554 | -0.052 | 0.523 | 0.413 | -0.072 | 0.528 | 0.460 | 0.071 | 0.045 | 0.310 | 0.475 |
| **S100A9** | -0.254 | 0.066 | 0.844 | 0.398 | 0.315 | 0.163 | -0.027 | -0.228 | 0.343 | -0.005 | 0.560 | -0.240 | 0.594 | 0.284 | -0.308 | 0.581 | 0.474 | -0.107 | -0.064 | 0.607 | 0.491 |
| **SET** | 0.039 | -0.140 | 0.480 | 0.380 | 0.053 | 0.186 | -0.359 | -0.327 | 0.169 | -0.196 | 0.319 | -0.309 | 0.436 | 0.153 | -0.324 | 0.574 | 0.315 | 0.194 | 0.000 | 0.661 | 0.353 |
| **SLC30A8** | -0.415 | -0.038 | 0.434 | -0.175 | -0.016 | 0.400 | 0.140 | 0.112 | 0.113 | 0.340 | 0.661 | -0.360 | 0.597 | 0.416 | 0.230 | 0.341 | 0.609 | 0.014 | -0.073 | 0.369 | 0.434 |
| **SNRNP70** | 0.429 | 0.108 | -0.260 | 1.066 | 0.556 | 0.069 | -0.143 | -0.053 | 0.424 | 0.222 | 0.414 | 0.005 | 0.502 | 0.511 | -0.436 | 0.568 | 0.363 | 0.260 | -0.189 | 0.140 | 0.487 |
| **SNRPA** | 0.564 | 0.025 | -0.225 | 0.786 | 0.112 | -0.397 | 0.039 | -0.120 | 0.214 | 0.069 | 0.169 | 0.234 | 0.408 | -0.181 | 0.040 | -0.098 | 0.145 | 0.118 | 0.239 | -0.188 | -0.284 |
| **SNRPB** | 0.834 | 0.549 | -0.389 | 1.357 | 0.171 | 0.325 | -0.052 | 0.379 | 0.546 | 0.466 | 0.403 | 0.654 | 0.537 | 0.461 | -0.725 | 0.538 | 0.424 | 0.733 | -0.344 | 0.039 | 0.544 |
| **SNRPD1** | -0.203 | -0.019 | -0.332 | 0.212 | -0.270 | -0.021 | -0.525 | 0.085 | -0.054 | -0.333 | 0.168 | 0.051 | 0.415 | 0.061 | -0.225 | 0.119 | 0.323 | 0.532 | 0.558 | 0.410 | 0.747 |
| **SOX13** | 0.255 | 0.108 | -0.321 | 0.154 | 0.509 | 0.065 | 0.106 | -0.056 | 0.182 | -0.126 | 0.711 | 0.244 | 0.441 | 0.174 | 0.341 | 0.148 | 0.790 | 0.265 | -0.057 | 0.172 | 0.698 |
| **SRP19** | 0.248 | 0.237 | 0.742 | 0.787 | 0.344 | 0.516 | 0.003 | 0.285 | 0.429 | 0.445 | 0.562 | 0.243 | 0.512 | 0.357 | 0.165 | 0.302 | 0.362 | 0.329 | -0.145 | 0.281 | 0.593 |
| **SSB** | 0.247 | 0.007 | -0.054 | 0.511 | -0.151 | -0.047 | -0.374 | 0.010 | 0.013 | 0.098 | 0.043 | 0.117 | 0.125 | -0.038 | -0.275 | 0.097 | 0.312 | 0.583 | 0.073 | 0.474 | 0.601 |
| **TG** | -0.112 | 0.141 | 0.527 | 0.353 | 0.343 | 0.391 | 0.005 | -0.121 | 0.364 | 0.078 | 0.378 | -0.496 | 0.242 | 0.249 | -0.203 | 0.567 | 0.155 | -0.113 | -0.163 | 0.685 | 0.481 |
| **TGFB1** | 0.288 | -0.365 | -0.157 | 0.499 | -0.087 | -0.231 | -0.403 | -0.280 | 0.135 | -0.222 | 0.041 | -0.060 | -0.102 | 0.247 | -1.042 | -0.160 | 0.189 | 0.092 | 0.812 | 0.867 | 0.520 |
| **TOP1** | -0.008 | 0.382 | -0.635 | 0.816 | 0.008 | 0.436 | -0.013 | 0.024 | 0.125 | 0.407 | 0.289 | 0.151 | 0.178 | 0.197 | -0.441 | 0.256 | 0.176 | 0.808 | 0.081 | 0.551 | 0.865 |
| **TRIM21** | -0.049 | 0.611 | 0.395 | 0.158 | 0.080 | 0.507 | 0.169 | 0.215 | 0.102 | 0.603 | 0.365 | 0.242 | 0.462 | -0.107 | -0.692 | 0.225 | 0.210 | 0.283 | -0.265 | 0.327 | 0.496 |
| **TROVE2** | -0.118 | 0.179 | 0.734 | 0.446 | 0.276 | 0.404 | -0.231 | -0.015 | 0.249 | 0.152 | 0.357 | -0.491 | 0.221 | 0.279 | -0.381 | 0.631 | 0.128 | -0.077 | 0.041 | 0.613 | 0.492 |
| **UBTF** | 0.051 | 0.143 | -0.408 | 0.775 | 0.025 | 0.195 | 0.040 | 0.013 | 0.088 | 0.164 | 0.407 | 0.099 | 0.296 | 0.240 | -0.124 | 0.303 | 0.355 | 0.409 | 0.011 | 0.464 | 0.769 |
| **VEGFA** | -0.236 | -0.063 | -0.681 | 0.665 | -0.091 | 0.162 | -0.033 | -0.232 | 0.197 | 0.157 | 0.554 | -0.014 | 0.657 | 0.113 | 0.123 | -0.035 | 0.365 | 0.259 | 0.519 | 0.754 | 0.824 |
| **VIM** | -0.191 | -0.380 | -0.126 | -0.090 | 0.337 | 0.123 | -0.010 | -0.102 | -0.124 | 0.070 | 0.490 | -0.234 | 0.150 | 0.029 | -0.298 | 0.448 | 0.065 | -0.229 | -0.264 | 0.212 | -0.022 |

**Table S4.** Associations of AABs reactivity with symptoms in men. P values from age-adjusted regression analysis comparing males with a specific symptom burden to males without the same symptom are shown. Last three columns show p values from age-adjusted regression analysis comparing males with different levels of symptoms burdens to the pre-pandemic healthy control group.

| **P value** | **Chest pain** | **Chills** | **Conjunctivitis** | **Cough productive** | **Diarrhea** | **Fatigue** | **Fever** | **Headache** | **Loss of appetite** | **Muscle aches** | **Nasal congestion** | **Nausea** | **Runny nose** | **Shortness of breath** | **Skin** | **Smell taste** | **Sneezing** | **Sore throat** | **Asymptomatic** | **Mild** | **More than mild** |
| --- | --- | --- | --- | --- | --- | --- | --- | --- | --- | --- | --- | --- | --- | --- | --- | --- | --- | --- | --- | --- | --- |
| **ACE2** | 0.154 | 0.462 | 0.719 | 0.068 | 0.088 | 0.399 | 0.737 | 0.559 | 0.360 | 0.061 | 0.004 | 0.966 | 0.023 | 0.043 | 0.623 | 0.036 | 0.096 | 0.208 | 0.222 | 0.140 | 0.137 |
| **AK4** | 0.677 | 0.573 | 0.173 | 0.024 | 0.208 | 0.148 | 0.946 | 0.426 | 0.352 | 0.370 | 0.064 | 0.664 | 0.395 | 0.424 | 0.811 | 0.377 | 0.371 | 0.307 | 0.431 | 0.980 | 0.699 |
| **ANXA2** | 0.574 | 0.260 | 0.808 | 0.043 | 0.313 | 0.613 | 0.730 | 0.648 | 0.125 | 0.681 | 0.050 | 0.577 | 0.316 | 0.220 | 0.594 | 0.086 | 0.182 | 0.405 | 0.731 | 0.122 | 0.129 |
| **APOH** | 0.151 | 0.738 | 0.256 | 0.089 | 0.586 | 0.779 | 0.960 | 0.823 | 0.448 | 0.492 | 0.827 | 0.055 | 0.616 | 0.573 | 0.998 | 0.048 | 0.352 | 0.044 | 0.036 | 0.153 | 0.068 |
| **AQP4** | 0.108 | 0.129 | 0.774 | 0.143 | 0.610 | 0.090 | 0.257 | 0.095 | 0.189 | 0.434 | 0.606 | 0.024 | 0.667 | 0.009 | 0.225 | 0.049 | 0.795 | 0.099 | 0.246 | 0.032 | 0.784 |
| **C3** | 0.794 | 0.692 | 0.310 | 0.164 | 0.157 | 0.197 | 0.717 | 0.945 | 0.163 | 0.663 | 0.085 | 0.251 | 0.244 | 0.209 | 0.707 | 0.022 | 0.321 | 0.994 | 0.503 | 0.032 | 0.150 |
| **CADM3** | 0.805 | 0.579 | 0.679 | 0.307 | 0.317 | 0.606 | 0.878 | 0.013 | 0.644 | 0.581 | 0.246 | 0.547 | 0.103 | 0.778 | 0.636 | 0.755 | 0.546 | 0.980 | 0.109 | 0.021 | 0.021 |
| **CHD4** | 0.656 | 0.902 | 0.602 | 0.003 | 0.369 | 0.566 | 0.720 | 0.619 | 0.156 | 0.990 | 0.008 | 0.978 | 0.085 | 0.664 | 0.400 | 0.241 | 0.020 | 0.039 | 0.494 | 0.124 | 0.085 |
| **CHGA** | 0.041 | 0.144 | 0.585 | 0.032 | 0.186 | 0.708 | 0.584 | 0.859 | 0.319 | 0.478 | 0.861 | 0.167 | 0.611 | 0.015 | 0.347 | 0.495 | 0.260 | 0.016 | 0.498 | 0.095 | 0.792 |
| **CSF2** | 0.120 | 0.598 | 0.716 | 0.034 | 0.018 | 0.495 | 0.852 | 0.340 | 0.453 | 0.053 | 0.033 | 0.304 | 0.104 | 0.143 | 0.451 | 0.108 | 0.208 | 0.197 | 0.182 | 0.500 | 0.314 |
| **CTLA4** | 0.453 | 0.571 | 0.613 | 0.250 | 0.517 | 0.483 | 0.612 | 0.451 | 0.536 | 0.955 | 0.018 | 0.351 | 0.073 | 0.132 | 0.680 | 0.013 | 0.160 | 0.783 | 0.598 | 0.012 | 0.108 |
| **CTSG** | 0.524 | 0.879 | 0.508 | 0.038 | 0.239 | 0.719 | 0.839 | 0.762 | 0.581 | 0.280 | 0.005 | 0.779 | 0.016 | 0.237 | 0.833 | 0.103 | 0.093 | 0.389 | 0.511 | 0.234 | 0.112 |
| **CXCL8** | 0.373 | 0.710 | 0.123 | 0.007 | 0.437 | 0.663 | 0.721 | 0.685 | 0.357 | 0.140 | 0.020 | 0.558 | 0.059 | 0.236 | 0.475 | 0.222 | 0.106 | 0.019 | 0.325 | 0.921 | 0.388 |
| **DBT** | 0.871 | 0.240 | 0.594 | 0.627 | 0.311 | 0.234 | 0.510 | 0.179 | 0.992 | 0.090 | 0.259 | 0.422 | 0.021 | 0.238 | 0.093 | 0.173 | 0.601 | 0.240 | 0.515 | 0.163 | 0.734 |
| **DLAT** | 0.817 | 0.638 | 0.644 | 0.494 | 0.198 | 0.680 | 0.355 | 0.618 | 0.995 | 0.620 | 0.228 | 0.323 | 0.354 | 0.997 | 0.320 | 0.040 | 0.326 | 0.808 | 0.417 | 0.178 | 0.096 |
| **ECE1** | 0.180 | 0.718 | 0.808 | <0.001 | 0.213 | 0.132 | 0.556 | 0.808 | 0.057 | 0.817 | 0.011 | 0.771 | 0.062 | 0.075 | 0.183 | 0.378 | 0.023 | 0.343 | 0.292 | 0.806 | 0.834 |
| **EIF4H** | 0.815 | 0.788 | 0.714 | 0.039 | 0.447 | 0.690 | 0.750 | 0.636 | 0.225 | 0.510 | 0.072 | 0.920 | 0.131 | 0.110 | 0.854 | 0.461 | 0.077 | 0.981 | 0.696 | 0.063 | 0.106 |
| **ELANE** | 0.615 | 0.518 | 0.330 | 0.372 | 0.693 | 0.380 | 0.993 | 0.691 | 0.592 | 0.331 | 0.029 | 0.607 | 0.064 | 0.418 | 0.353 | 0.031 | 0.184 | 0.893 | 0.858 | 0.185 | 0.046 |
| **EXOSC10** | 0.854 | 0.325 | 0.248 | 0.013 | 0.676 | 0.508 | 0.660 | 0.920 | 0.787 | 0.514 | 0.547 | 0.906 | 0.572 | 0.572 | 0.496 | 0.145 | 0.348 | 0.212 | 0.532 | 0.106 | 0.305 |
| **GRP** | 0.173 | 0.203 | 0.947 | 0.001 | 0.194 | 0.423 | 0.425 | 0.157 | 0.192 | 0.421 | 0.378 | 0.041 | 0.427 | 0.443 | 0.584 | 0.717 | 0.213 | 0.064 | 0.185 | 0.614 | 0.966 |
| **HARS** | 0.854 | 0.577 | 0.427 | 0.053 | 0.301 | 0.138 | 0.787 | 0.782 | 0.341 | 0.543 | 0.047 | 0.292 | 0.269 | 0.269 | 0.631 | 0.047 | 0.210 | 0.496 | 0.907 | 0.007 | 0.047 |
| **HIST1H4A** | 0.748 | 0.369 | 0.431 | 0.003 | 0.506 | 0.253 | 0.591 | 0.932 | 0.704 | 0.218 | 0.020 | 0.591 | 0.024 | 0.237 | 0.435 | 0.192 | 0.199 | 0.049 | 0.567 | 0.713 | 0.479 |
| **ICA1** | 0.986 | 0.899 | 0.117 | 0.009 | 0.510 | 0.283 | 0.947 | 0.409 | 0.648 | 0.453 | 0.143 | 0.311 | 0.954 | 0.132 | 0.926 | 0.276 | 0.210 | 0.178 | 0.677 | 0.898 | 0.122 |
| **IFNA4** | 0.845 | 0.899 | 0.303 | 0.150 | 0.298 | 0.161 | 0.978 | 0.896 | 0.246 | 0.552 | 0.011 | 0.364 | 0.032 | 0.285 | 0.548 | 0.014 | 0.081 | 0.901 | 0.346 | 0.023 | 0.107 |
| **IFNA6** | 0.399 | 0.597 | 0.403 | 0.045 | 0.432 | 0.213 | 0.641 | 0.853 | 0.440 | 0.162 | 0.010 | 0.994 | 0.008 | 0.214 | 0.883 | 0.035 | 0.103 | 0.312 | 0.147 | 0.666 | 0.591 |
| **IGF1R** | 0.881 | 0.739 | 0.544 | 0.394 | 0.108 | 0.454 | 0.955 | 0.918 | 0.479 | 0.407 | 0.053 | 0.247 | 0.245 | 0.357 | 0.593 | 0.014 | 0.499 | 0.664 | 0.440 | 0.107 | 0.201 |
| **IL17A** | 0.506 | 0.621 | 0.389 | 0.014 | 0.054 | 0.765 | 0.732 | 0.882 | 0.039 | 0.874 | 0.067 | 0.031 | 0.864 | 0.070 | 0.619 | 0.755 | 0.134 | 0.301 | 0.928 | 0.453 | 0.744 |
| **IL4** | 0.488 | 0.793 | 0.835 | 0.115 | 0.066 | 0.798 | 0.966 | 0.279 | 0.123 | 0.351 | 0.016 | 0.900 | 0.077 | 0.304 | 0.550 | 0.470 | 0.015 | 0.776 | 0.607 | 0.477 | 0.248 |
| **IL6** | 0.469 | 0.716 | 0.937 | 0.099 | 0.719 | 0.489 | 0.782 | 0.028 | 0.861 | 0.219 | 0.096 | 0.395 | 0.392 | 0.430 | 0.642 | 0.661 | 0.154 | 0.637 | 0.588 | 0.014 | 0.262 |
| **INS** | 0.224 | 0.218 | 0.482 | 0.005 | 0.232 | 0.795 | 0.252 | 0.163 | 0.679 | 0.399 | 0.323 | 0.792 | 0.250 | 0.929 | 0.742 | 0.918 | 0.251 | 0.620 | 0.040 | 0.001 | 0.072 |
| **LYZ** | 0.707 | 0.843 | 0.360 | 0.058 | 0.205 | 0.331 | 0.951 | 0.776 | 0.422 | 0.192 | 0.001 | 0.833 | 0.006 | 0.120 | 0.970 | 0.033 | 0.032 | 0.524 | 0.206 | 0.172 | 0.128 |
| **MDA5** | 0.444 | 0.983 | 0.771 | <0.001 | 0.309 | 0.741 | 0.447 | 0.782 | 0.252 | 0.933 | 0.015 | 0.143 | 0.095 | 0.350 | 0.379 | 0.186 | 0.096 | 0.005 | 0.697 | 0.044 | 0.038 |
| **MIF** | 0.969 | 0.478 | 0.640 | 0.003 | 0.703 | 0.344 | 0.977 | 0.656 | 0.489 | 0.369 | 0.065 | 0.229 | 0.073 | 0.347 | 0.900 | 0.329 | 0.070 | 0.008 | 0.383 | 0.006 | 0.001 |
| **MOV10** | 0.613 | 0.859 | 0.073 | 0.058 | 0.019 | 0.306 | 0.397 | 0.918 | 0.040 | 0.208 | 0.015 | 0.161 | 0.200 | 0.216 | 0.249 | 0.333 | 0.079 | 0.047 | 0.330 | 0.113 | 0.039 |
| **MX1** | 0.788 | 0.993 | 0.245 | 0.003 | 0.712 | 0.763 | 0.902 | 0.099 | 0.973 | 0.987 | 0.027 | 0.910 | 0.061 | 0.306 | 0.428 | 0.148 | 0.119 | 0.461 | 0.391 | 0.016 | 0.020 |
| **NCL** | 0.856 | 0.437 | 0.190 | 0.004 | 0.292 | 0.174 | 0.439 | 0.853 | 0.104 | 0.320 | 0.020 | 0.266 | 0.421 | 0.059 | 0.377 | 0.265 | 0.279 | 0.026 | 0.805 | 0.126 | 0.015 |
| **NPM1** | 0.661 | 0.363 | 0.874 | 0.026 | 0.723 | 0.346 | 0.457 | 0.399 | 0.548 | 0.632 | 0.128 | 0.493 | 0.122 | 0.581 | 0.719 | 0.036 | 0.197 | 0.928 | 0.940 | 0.007 | 0.073 |
| **PRTN3** | 0.742 | 0.557 | 0.666 | 0.164 | 0.917 | 0.770 | 0.796 | 0.101 | 0.838 | 0.409 | 0.013 | 0.738 | 0.046 | 0.284 | 0.914 | 0.606 | 0.014 | 0.697 | 0.594 | 0.246 | 0.292 |
| **RAE1** | 0.466 | 0.025 | 0.043 | 0.507 | 0.169 | 0.220 | 0.325 | 0.488 | 0.180 | 0.450 | 0.101 | 0.290 | 0.094 | 0.798 | 0.658 | 0.500 | 0.151 | 0.690 | 0.197 | 0.206 | 0.230 |
| **RNF41** | 0.956 | 0.803 | 0.736 | 0.035 | 0.640 | 0.992 | 0.965 | 0.405 | 0.478 | 0.892 | 0.025 | 0.809 | 0.034 | 0.217 | 0.353 | 0.043 | 0.085 | 0.669 | 0.450 | 0.012 | 0.024 |
| **RPLP2** | 0.952 | 0.874 | 0.484 | 0.146 | 0.323 | 0.408 | 0.894 | 0.533 | 0.217 | 0.484 | 0.036 | 0.886 | 0.068 | 0.128 | 0.894 | 0.045 | 0.091 | 0.796 | 0.900 | 0.160 | 0.139 |
| **S100A9** | 0.448 | 0.813 | 0.065 | 0.241 | 0.255 | 0.560 | 0.923 | 0.416 | 0.231 | 0.984 | 0.034 | 0.508 | 0.037 | 0.298 | 0.567 | 0.027 | 0.081 | 0.696 | 0.847 | 0.009 | 0.093 |
| **SET** | 0.899 | 0.589 | 0.261 | 0.226 | 0.838 | 0.472 | 0.155 | 0.205 | 0.524 | 0.428 | 0.197 | 0.356 | 0.102 | 0.545 | 0.516 | 0.018 | 0.214 | 0.445 | 1.000 | 0.006 | 0.208 |
| **SLC30A8** | 0.169 | 0.880 | 0.298 | 0.569 | 0.950 | 0.111 | 0.573 | 0.659 | 0.664 | 0.158 | 0.005 | 0.270 | 0.020 | 0.089 | 0.637 | 0.154 | 0.012 | 0.954 | 0.831 | 0.176 | 0.117 |
| **SNRNP70** | 0.281 | 0.746 | 0.638 | 0.007 | 0.090 | 0.837 | 0.663 | 0.874 | 0.213 | 0.486 | 0.192 | 0.990 | 0.144 | 0.113 | 0.497 | 0.071 | 0.266 | 0.426 | 0.697 | 0.611 | 0.258 |
| **SNRPA** | 0.103 | 0.932 | 0.640 | 0.024 | 0.699 | 0.170 | 0.893 | 0.680 | 0.473 | 0.804 | 0.544 | 0.534 | 0.174 | 0.526 | 0.943 | 0.724 | 0.611 | 0.680 | 0.613 | 0.508 | 0.404 |
| **SNRPB** | 0.005 | 0.028 | 0.353 | <0.001 | 0.498 | 0.198 | 0.833 | 0.133 | 0.033 | 0.051 | 0.093 | 0.043 | 0.038 | 0.060 | 0.135 | 0.023 | 0.085 | 0.002 | 0.383 | 0.877 | 0.110 |
| **SNRPD1** | 0.536 | 0.945 | 0.462 | 0.523 | 0.319 | 0.940 | 0.047 | 0.755 | 0.848 | 0.201 | 0.519 | 0.886 | 0.140 | 0.820 | 0.669 | 0.648 | 0.225 | 0.043 | 0.078 | 0.066 | 0.022 |
| **SOX13** | 0.399 | 0.670 | 0.443 | 0.616 | 0.040 | 0.797 | 0.669 | 0.826 | 0.483 | 0.603 | 0.002 | 0.456 | 0.089 | 0.481 | 0.484 | 0.539 | 0.001 | 0.283 | 0.888 | 0.540 | 0.020 |
| **SRP19** | 0.433 | 0.370 | 0.086 | 0.012 | 0.187 | 0.047 | 0.992 | 0.280 | 0.110 | 0.075 | 0.023 | 0.478 | 0.059 | 0.164 | 0.746 | 0.228 | 0.160 | 0.202 | 0.720 | 0.270 | 0.033 |
| **SSB** | 0.415 | 0.978 | 0.897 | 0.093 | 0.547 | 0.851 | 0.129 | 0.967 | 0.961 | 0.686 | 0.860 | 0.722 | 0.635 | 0.878 | 0.573 | 0.687 | 0.207 | 0.016 | 0.852 | 0.098 | 0.027 |
| **TG** | 0.747 | 0.625 | 0.267 | 0.313 | 0.229 | 0.172 | 0.985 | 0.675 | 0.216 | 0.777 | 0.168 | 0.181 | 0.417 | 0.376 | 0.715 | 0.036 | 0.585 | 0.688 | 0.607 | 0.006 | 0.127 |
| **TGFB1** | 0.381 | 0.181 | 0.730 | 0.132 | 0.751 | 0.400 | 0.131 | 0.307 | 0.633 | 0.398 | 0.876 | 0.867 | 0.720 | 0.356 | 0.045 | 0.541 | 0.483 | 0.733 | 0.051 | 0.008 | 0.056 |
| **TOP1** | 0.980 | 0.134 | 0.132 | 0.007 | 0.974 | 0.086 | 0.958 | 0.925 | 0.636 | 0.095 | 0.238 | 0.651 | 0.505 | 0.433 | 0.372 | 0.295 | 0.484 | 0.001 | 0.815 | 0.029 | 0.001 |
| **TRIM21** | 0.885 | 0.028 | 0.399 | 0.647 | 0.775 | 0.070 | 0.543 | 0.447 | 0.727 | 0.023 | 0.176 | 0.509 | 0.113 | 0.698 | 0.203 | 0.405 | 0.450 | 0.307 | 0.319 | 0.241 | 0.073 |
| **TROVE2** | 0.738 | 0.544 | 0.129 | 0.211 | 0.345 | 0.167 | 0.423 | 0.960 | 0.410 | 0.589 | 0.203 | 0.196 | 0.469 | 0.331 | 0.502 | 0.022 | 0.658 | 0.791 | 0.893 | 0.010 | 0.108 |
| **UBTF** | 0.847 | 0.517 | 0.261 | 0.003 | 0.908 | 0.376 | 0.852 | 0.954 | 0.696 | 0.436 | 0.050 | 0.730 | 0.192 | 0.262 | 0.770 | 0.146 | 0.097 | 0.055 | 0.979 | 0.059 | 0.005 |
| **VEGFA** | 0.459 | 0.813 | 0.119 | 0.037 | 0.731 | 0.543 | 0.901 | 0.382 | 0.470 | 0.537 | 0.027 | 0.968 | 0.015 | 0.665 | 0.811 | 0.890 | 0.160 | 0.319 | 0.108 | 0.008 | 0.001 |
| **VIM** | 0.524 | 0.125 | 0.761 | 0.768 | 0.172 | 0.622 | 0.967 | 0.683 | 0.630 | 0.769 | 0.037 | 0.469 | 0.564 | 0.906 | 0.536 | 0.057 | 0.790 | 0.348 | 0.539 | 0.529 | 0.942 |

**Table S5.** Associations of AABs reactivity with symptoms in women. Beta coefficients from age-adjusted regression analysis comparing males with a specific symptom burden to females without the same symptom are shown. Last three columns show beta coefficients from age-adjusted regression analysis comparing females with different levels of symptoms burdens to the pre-pandemic healthy control group.

| **Beta** | **Chest pain** | **Chills** | **Conjunctivitis** | **Cough dry** | **Fever** | **Loss of appetite** | **Nasal congestion** | **Nausea** | **Shortness of breath** | **Skin** | **Smell taste** | **Sore throat** | **Vomiting** | **Asymptomatic** | **Mild** | **More than mild** |
| --- | --- | --- | --- | --- | --- | --- | --- | --- | --- | --- | --- | --- | --- | --- | --- | --- |
| **AQP4** | -0.093 | 0.029 | -0.029 | -0.150 | 0.130 | -0.174 | -0.251 | -0.262 | -0.022 | -0.252 | -0.201 | -0.465 | 0.207 | 0.479 | 0.499 | -0.121 |
| **C3** | -0.388 | -0.248 | -0.372 | -0.130 | 0.091 | -0.158 | -0.138 | -0.048 | -0.064 | 0.162 | 0.177 | -0.173 | 0.470 | 0.429 | 0.492 | 0.274 |
| **CENPB** | -0.065 | -0.169 | -0.328 | -0.239 | 0.036 | -0.141 | -0.046 | 0.069 | 0.161 | 0.004 | 0.204 | -0.043 | 0.700 | 0.369 | 0.357 | 0.443 |
| **CHD3** | -0.385 | -0.438 | 0.019 | -0.102 | -0.260 | -0.117 | -0.266 | -0.021 | -0.007 | 0.796 | 0.124 | -0.305 | 0.200 | 1.041 | 1.062 | 0.485 |
| **CHD4** | 0.054 | -0.404 | -0.691 | -0.203 | -0.116 | -0.280 | 0.120 | 0.028 | 0.244 | 0.153 | 0.180 | -0.156 | -0.007 | 0.111 | 0.051 | 0.298 |
| **CHGA** | 0.052 | -0.149 | 0.015 | -0.003 | 0.096 | -0.118 | -0.349 | -0.014 | 0.525 | -0.471 | -0.046 | -0.187 | 0.062 | 0.215 | 0.007 | 0.068 |
| **DBT** | -0.333 | -0.448 | -0.426 | -0.400 | -0.439 | 0.081 | 0.096 | -0.038 | 0.001 | 0.033 | 0.032 | 0.055 | -0.100 | 0.740 | 0.765 | 0.479 |
| **ECE1** | 0.069 | -0.178 | -0.044 | -0.285 | 0.055 | -0.252 | 0.082 | 0.054 | 0.240 | -0.288 | 0.345 | -0.125 | 0.737 | 0.689 | 0.478 | 0.313 |
| **ELANE** | 0.351 | -0.113 | 1.044 | -0.245 | -0.166 | -0.502 | -0.094 | 0.140 | 0.167 | -0.065 | -0.104 | -0.066 | 0.105 | 0.548 | 0.305 | 0.332 |
| **EXOSC10** | -0.029 | -0.222 | -0.464 | -0.429 | -0.187 | -0.471 | -0.240 | -0.031 | 0.108 | 0.078 | -0.115 | -0.150 | 0.195 | 0.992 | 0.372 | 0.396 |
| **GAD65** | -0.246 | -0.193 | -0.248 | -0.275 | -0.098 | -0.415 | 0.011 | -0.157 | 0.090 | -0.320 | 0.129 | -0.162 | -0.059 | 0.185 | 0.211 | -0.113 |
| **HARS** | -0.108 | -0.215 | 0.042 | -0.197 | 0.116 | -0.059 | -0.175 | 0.085 | 0.025 | -0.313 | 0.223 | -0.178 | 0.697 | 0.380 | 0.416 | 0.338 |
| **HIST1H4A** | -0.071 | -0.129 | -0.315 | -0.399 | -0.088 | -0.267 | -0.109 | -0.019 | 0.257 | 0.083 | 0.052 | -0.107 | 0.326 | 0.419 | 0.540 | 0.399 |
| **IFNA2** | -0.404 | -0.199 | -0.556 | -0.191 | 0.193 | -0.103 | -0.238 | -0.046 | 0.058 | -0.127 | -0.156 | 0.138 | 0.632 | 0.421 | 0.376 | 0.042 |
| **IGF1R** | -0.437 | -0.124 | -0.558 | -0.097 | 0.031 | -0.185 | -0.218 | -0.102 | -0.207 | -0.179 | -0.120 | -0.170 | 0.445 | 0.471 | 0.629 | 0.303 |
| **IL10** | -0.230 | -0.330 | -0.160 | -0.366 | -0.155 | -0.248 | -0.124 | -0.011 | 0.000 | 0.050 | 0.001 | -0.217 | 0.247 | 0.760 | 0.277 | 0.293 |
| **INS** | 0.310 | -0.112 | 1.450 | 0.012 | -0.211 | -0.251 | -0.079 | -0.002 | -0.172 | -0.454 | 0.048 | -0.062 | 0.238 | 0.442 | 0.510 | 0.428 |
| **MOV10** | -0.150 | 0.022 | 1.161 | -0.025 | 0.000 | -0.024 | -0.159 | -0.128 | -0.208 | 0.249 | 0.149 | 0.069 | 0.087 | -0.195 | -0.063 | -0.161 |
| **MX1** | 0.076 | -0.243 | -0.525 | -0.253 | -0.227 | -0.500 | -0.139 | -0.040 | 0.262 | 0.079 | 0.113 | -0.104 | 0.183 | 0.741 | 0.524 | 0.468 |
| **PRTN3** | 0.273 | -0.201 | 0.105 | -0.144 | 0.000 | -0.475 | 0.181 | -0.227 | 0.285 | 0.987 | -0.233 | -0.340 | -0.158 | 0.384 | 0.574 | 0.277 |
| **PTPRN** | 0.227 | 0.072 | -0.083 | -0.036 | 0.357 | 0.024 | 0.170 | 0.034 | 0.299 | -0.221 | 0.311 | 0.053 | 0.086 | -0.288 | 0.040 | 0.151 |
| **RNF41** | -0.140 | -0.129 | -0.544 | -0.233 | 0.031 | -0.423 | -0.091 | -0.096 | 0.091 | -0.250 | 0.121 | -0.134 | 0.296 | 0.677 | 0.295 | 0.486 |
| **ROS1** | -0.094 | -0.051 | -0.314 | -0.214 | -0.067 | -0.371 | -0.394 | -0.124 | -0.144 | -0.318 | -0.380 | -0.262 | 0.172 | 0.395 | 0.203 | 0.036 |
| **RPLP2** | -0.234 | -0.243 | -0.269 | -0.264 | 0.048 | -0.160 | -0.249 | -0.044 | 0.047 | -0.221 | 0.060 | -0.053 | 0.624 | 0.292 | 0.447 | 0.025 |
| **S100A8** | -0.161 | 0.219 | 0.547 | -0.077 | 0.383 | 0.189 | -0.129 | -0.176 | 0.142 | -0.480 | 0.063 | -0.028 | 0.289 | 0.504 | 0.178 | 0.472 |
| **S100A9** | -0.355 | -0.353 | -0.214 | -0.335 | -0.149 | -0.373 | -0.286 | -0.088 | -0.004 | -0.236 | 0.060 | -0.298 | 0.309 | 1.036 | 0.544 | 0.356 |
| **SET** | -0.046 | -0.248 | -0.482 | -0.327 | -0.121 | -0.527 | -0.043 | -0.232 | -0.029 | 0.224 | -0.254 | -0.214 | 0.424 | 0.553 | 0.303 | 0.038 |
| **SMD3** | 0.041 | -0.288 | -0.175 | -0.417 | -0.301 | -0.269 | -0.004 | 0.027 | 0.096 | 0.152 | 0.270 | -0.005 | 0.259 | 0.325 | 0.316 | 0.287 |
| **SNRPC** | 0.124 | -0.130 | 0.071 | -0.075 | 0.148 | -0.021 | -0.049 | 0.393 | -0.207 | -0.196 | -0.022 | 0.237 | 0.306 | 0.136 | 0.185 | 0.465 |
| **SOX13** | 0.088 | -0.299 | -0.182 | -0.155 | -0.018 | -0.028 | 0.219 | 0.039 | 0.269 | -0.701 | -0.179 | -0.034 | -0.475 | 0.701 | 1.094 | 0.693 |
| **SP100** | 0.140 | 0.080 | -0.023 | -0.015 | 0.249 | -0.102 | -0.040 | 0.016 | 0.530 | -0.203 | -0.063 | 0.208 | 0.706 | 0.402 | 0.172 | 0.759 |
| **SRP54** | -0.485 | -0.188 | 0.225 | -0.084 | -0.076 | 0.120 | -0.119 | -0.140 | -0.130 | 0.004 | -0.005 | 0.011 | -0.322 | 0.219 | 0.539 | 0.185 |
| **TG** | -0.449 | -0.256 | -0.405 | -0.144 | 0.104 | -0.131 | -0.251 | 0.015 | -0.117 | -0.182 | 0.151 | -0.174 | 0.527 | 0.506 | 0.479 | 0.268 |
| **TGFB1** | 0.258 | -0.142 | -0.090 | -0.298 | -0.286 | -0.250 | -0.068 | 0.005 | 0.191 | 0.072 | 0.079 | -0.382 | 0.190 | -0.077 | -0.218 | -0.118 |
| **TOP1** | -0.088 | -0.200 | -0.297 | -0.476 | -0.183 | -0.390 | -0.046 | -0.001 | 0.111 | 0.094 | 0.179 | -0.201 | 0.302 | 0.462 | 0.421 | 0.344 |
| **TPO** | -0.410 | -0.098 | -0.621 | -0.043 | 0.174 | -0.063 | -0.004 | -0.434 | 0.072 | 0.148 | 0.036 | -0.506 | 0.128 | 0.553 | 0.608 | 0.159 |
| **TRIM33** | 0.024 | -0.233 | 0.267 | -0.315 | -0.104 | -0.177 | -0.018 | 0.098 | 0.100 | 0.201 | 0.004 | -0.119 | -0.086 | -0.004 | -0.025 | -0.194 |
| **UBTF** | -0.031 | -0.221 | -0.209 | -0.503 | -0.209 | -0.228 | -0.141 | 0.016 | 0.090 | 0.160 | 0.283 | -0.211 | 0.281 | 0.580 | 0.394 | 0.126 |

**Table S6.** Associations of AABs reactivity with symptoms in women. P values from age-adjusted regression analysis comparing males with a specific symptom burden to females without the same symptom are shown. Last three columns show p values from age-adjusted regression analysis comparing females with different levels of symptoms burdens to the pre-pandemic healthy control group.

| **P value** | **Chest pain** | **Chills** | **Conjunctivitis** | **Cough dry** | **Fever** | **Loss of appetite** | **Nasal congestion** | **Nausea** | **Shortness of breath** | **Skin** | **Smell taste** | **Sore throat** | **Vomiting** | **Asymptomatic** | **Mild** | **More than mild** |
| --- | --- | --- | --- | --- | --- | --- | --- | --- | --- | --- | --- | --- | --- | --- | --- | --- |
| **AQP4** | 0.690 | 0.879 | 0.951 | 0.429 | 0.502 | 0.363 | 0.183 | 0.193 | 0.918 | 0.473 | 0.289 | 0.014 | 0.556 | 0.260 | 0.154 | 0.668 |
| **C3** | 0.047 | 0.121 | 0.346 | 0.418 | 0.578 | 0.327 | 0.385 | 0.776 | 0.714 | 0.585 | 0.268 | 0.285 | 0.111 | 0.059 | 0.075 | 0.222 |
| **CENPB** | 0.773 | 0.353 | 0.466 | 0.189 | 0.845 | 0.444 | 0.800 | 0.721 | 0.421 | 0.991 | 0.260 | 0.814 | 0.036 | 0.132 | 0.200 | 0.096 |
| **CHD3** | 0.106 | 0.023 | 0.968 | 0.600 | 0.190 | 0.553 | 0.170 | 0.921 | 0.975 | 0.026 | 0.525 | 0.118 | 0.580 | 0.003 | 0.003 | 0.110 |
| **CHD4** | 0.822 | 0.038 | 0.152 | 0.299 | 0.562 | 0.156 | 0.539 | 0.893 | 0.256 | 0.675 | 0.357 | 0.430 | 0.984 | 0.751 | 0.882 | 0.340 |
| **CHGA** | 0.822 | 0.434 | 0.974 | 0.989 | 0.619 | 0.540 | 0.064 | 0.944 | 0.011 | 0.179 | 0.810 | 0.329 | 0.859 | 0.651 | 0.984 | 0.810 |
| **DBT** | 0.134 | 0.012 | 0.340 | 0.026 | 0.016 | 0.660 | 0.594 | 0.844 | 0.996 | 0.922 | 0.859 | 0.762 | 0.765 | 0.075 | 0.012 | 0.113 |
| **ECE1** | 0.739 | 0.294 | 0.916 | 0.090 | 0.750 | 0.139 | 0.629 | 0.763 | 0.196 | 0.358 | 0.039 | 0.466 | 0.017 | 0.016 | 0.111 | 0.184 |
| **ELANE** | 0.141 | 0.563 | 0.028 | 0.207 | 0.402 | 0.010 | 0.627 | 0.500 | 0.435 | 0.856 | 0.592 | 0.735 | 0.770 | 0.044 | 0.352 | 0.191 |
| **EXOSC10** | 0.906 | 0.266 | 0.347 | 0.030 | 0.359 | 0.018 | 0.227 | 0.883 | 0.624 | 0.834 | 0.563 | 0.458 | 0.598 | 0.002 | 0.255 | 0.184 |
| **GAD65** | 0.249 | 0.267 | 0.563 | 0.111 | 0.579 | 0.017 | 0.948 | 0.395 | 0.638 | 0.320 | 0.456 | 0.354 | 0.855 | 0.632 | 0.491 | 0.614 |
| **HARS** | 0.637 | 0.247 | 0.926 | 0.290 | 0.539 | 0.756 | 0.346 | 0.668 | 0.902 | 0.363 | 0.229 | 0.342 | 0.041 | 0.165 | 0.162 | 0.230 |
| **HIST1H4A** | 0.779 | 0.526 | 0.531 | 0.049 | 0.672 | 0.193 | 0.592 | 0.931 | 0.249 | 0.827 | 0.797 | 0.604 | 0.387 | 0.176 | 0.088 | 0.165 |
| **IFNA2** | 0.052 | 0.243 | 0.185 | 0.262 | 0.265 | 0.551 | 0.160 | 0.799 | 0.756 | 0.688 | 0.358 | 0.422 | 0.043 | 0.099 | 0.191 | 0.862 |
| **IGF1R** | 0.038 | 0.472 | 0.189 | 0.573 | 0.860 | 0.287 | 0.203 | 0.579 | 0.273 | 0.575 | 0.485 | 0.327 | 0.161 | 0.139 | 0.052 | 0.239 |
| **IL10** | 0.295 | 0.063 | 0.716 | 0.039 | 0.396 | 0.169 | 0.487 | 0.955 | 1.000 | 0.879 | 0.995 | 0.227 | 0.455 | 0.022 | 0.397 | 0.288 |
| **INS** | 0.221 | 0.587 | 0.004 | 0.954 | 0.315 | 0.228 | 0.702 | 0.993 | 0.449 | 0.234 | 0.817 | 0.765 | 0.533 | 0.179 | 0.155 | 0.123 |
| **MOV10** | 0.446 | 0.890 | 0.003 | 0.878 | 0.999 | 0.883 | 0.317 | 0.452 | 0.234 | 0.398 | 0.350 | 0.666 | 0.769 | 0.585 | 0.840 | 0.531 |
| **MX1** | 0.750 | 0.207 | 0.270 | 0.188 | 0.247 | 0.009 | 0.469 | 0.847 | 0.215 | 0.826 | 0.558 | 0.595 | 0.608 | 0.012 | 0.103 | 0.069 |
| **PRTN3** | 0.271 | 0.319 | 0.833 | 0.476 | 0.998 | 0.019 | 0.369 | 0.289 | 0.197 | 0.007 | 0.247 | 0.094 | 0.673 | 0.344 | 0.151 | 0.366 |
| **PTPRN** | 0.298 | 0.684 | 0.850 | 0.837 | 0.046 | 0.893 | 0.334 | 0.857 | 0.123 | 0.501 | 0.076 | 0.769 | 0.793 | 0.427 | 0.906 | 0.615 |
| **RNF41** | 0.569 | 0.518 | 0.270 | 0.243 | 0.878 | 0.035 | 0.650 | 0.651 | 0.680 | 0.499 | 0.544 | 0.508 | 0.424 | 0.034 | 0.356 | 0.069 |
| **ROS1** | 0.651 | 0.764 | 0.449 | 0.202 | 0.698 | 0.027 | 0.017 | 0.488 | 0.434 | 0.306 | 0.022 | 0.121 | 0.581 | 0.261 | 0.507 | 0.894 |
| **RPLP2** | 0.235 | 0.128 | 0.497 | 0.098 | 0.768 | 0.323 | 0.118 | 0.798 | 0.789 | 0.456 | 0.710 | 0.746 | 0.034 | 0.461 | 0.184 | 0.926 |
| **S100A8** | 0.475 | 0.232 | 0.226 | 0.677 | 0.039 | 0.309 | 0.483 | 0.368 | 0.482 | 0.157 | 0.731 | 0.881 | 0.395 | 0.226 | 0.579 | 0.116 |
| **S100A9** | 0.115 | 0.054 | 0.638 | 0.067 | 0.426 | 0.043 | 0.117 | 0.655 | 0.984 | 0.488 | 0.745 | 0.107 | 0.363 | 0.001 | 0.072 | 0.174 |
| **SET** | 0.839 | 0.177 | 0.289 | 0.073 | 0.518 | 0.004 | 0.813 | 0.236 | 0.885 | 0.511 | 0.165 | 0.248 | 0.212 | 0.178 | 0.378 | 0.896 |
| **SMD3** | 0.861 | 0.131 | 0.712 | 0.028 | 0.121 | 0.163 | 0.984 | 0.894 | 0.646 | 0.668 | 0.155 | 0.979 | 0.465 | 0.239 | 0.312 | 0.277 |
| **SNRPC** | 0.570 | 0.464 | 0.872 | 0.674 | 0.415 | 0.907 | 0.784 | 0.036 | 0.289 | 0.551 | 0.902 | 0.185 | 0.352 | 0.689 | 0.540 | 0.095 |
| **SOX13** | 0.693 | 0.098 | 0.685 | 0.392 | 0.923 | 0.879 | 0.224 | 0.840 | 0.175 | 0.035 | 0.321 | 0.851 | 0.155 | 0.072 | 0.001 | 0.014 |
| **SP100** | 0.572 | 0.692 | 0.964 | 0.943 | 0.226 | 0.618 | 0.843 | 0.939 | 0.016 | 0.586 | 0.756 | 0.307 | 0.057 | 0.103 | 0.532 | 0.016 |
| **SRP54** | 0.018 | 0.263 | 0.589 | 0.620 | 0.660 | 0.481 | 0.480 | 0.434 | 0.484 | 0.990 | 0.975 | 0.950 | 0.302 | 0.611 | 0.138 | 0.517 |
| **TG** | 0.031 | 0.131 | 0.336 | 0.398 | 0.550 | 0.446 | 0.138 | 0.936 | 0.531 | 0.564 | 0.375 | 0.312 | 0.093 | 0.040 | 0.097 | 0.274 |
| **TGFB1** | 0.231 | 0.419 | 0.835 | 0.088 | 0.107 | 0.156 | 0.698 | 0.979 | 0.320 | 0.825 | 0.651 | 0.029 | 0.559 | 0.849 | 0.574 | 0.689 |
| **TOP1** | 0.726 | 0.326 | 0.555 | 0.018 | 0.379 | 0.056 | 0.823 | 0.998 | 0.620 | 0.803 | 0.380 | 0.329 | 0.423 | 0.150 | 0.221 | 0.247 |
| **TPO** | 0.081 | 0.612 | 0.190 | 0.822 | 0.375 | 0.747 | 0.984 | 0.032 | 0.732 | 0.677 | 0.850 | 0.008 | 0.720 | 0.107 | 0.082 | 0.579 |
| **TRIM33** | 0.902 | 0.144 | 0.499 | 0.047 | 0.525 | 0.273 | 0.912 | 0.564 | 0.568 | 0.497 | 0.979 | 0.462 | 0.771 | 0.993 | 0.942 | 0.487 |
| **UBTF** | 0.908 | 0.303 | 0.692 | 0.018 | 0.337 | 0.292 | 0.509 | 0.944 | 0.703 | 0.686 | 0.185 | 0.328 | 0.478 | 0.057 | 0.295 | 0.654 |

**Figure S1. Sex-specific associations of autoantibodies with SLE status.** In age-adjusted regression analyses, the breadth and magnitude of associations observed AABs reactivity and systemic lupus erythematosus (SLE) compared to health control status were predominantly seen in women compared to men. Beta coefficients were shown to the left, and negative log P values were shown to the right in each panel.

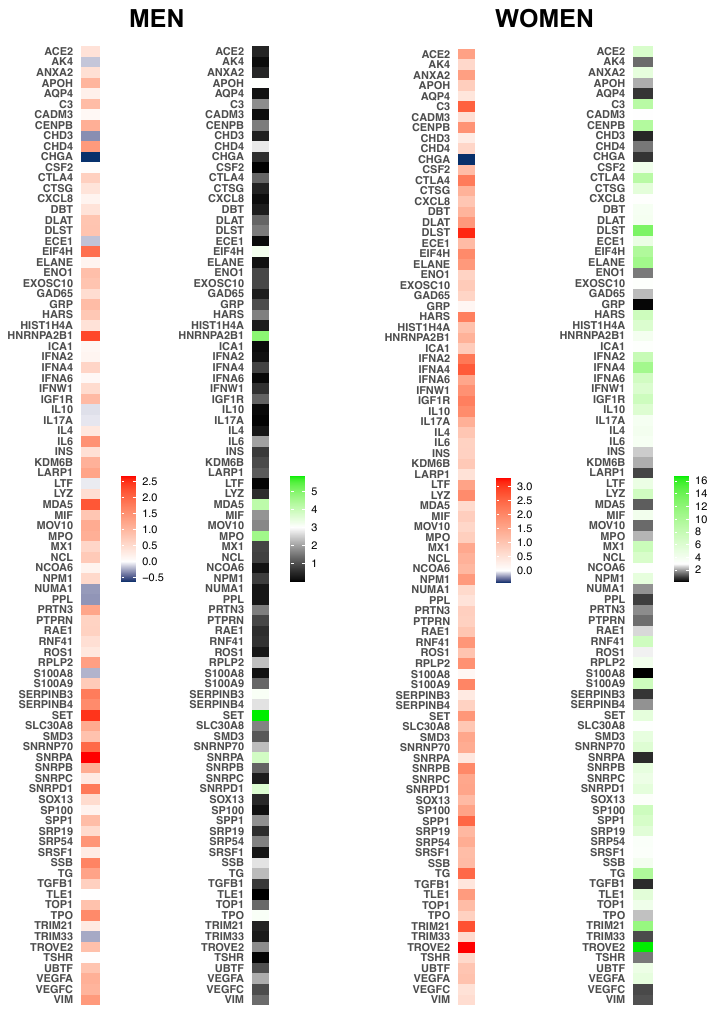
